# Identification of atypical circulating tumor cells with prognostic value in metastatic breast cancer patients

**DOI:** 10.1101/2020.09.29.20203653

**Authors:** Alexia Lopresti, Laurys Boudin, Pascal Finetti, Séverine Garnier, Anaïs Aulas, Maria-Lucia Liberatoscioli, Olivier Cabaud, Arnaud Guille, Alexandre de Nonneville, Quentin Dacosta, Emilie Denicolai, Jihane Pakradouni, Anthony Goncalves, Daniel Birnbaum, Claire Acquaviva, François Bertucci, Emilie Mamessier

## Abstract

**Background:** Circulating tumor cells (CTCs) have a strong potential as a quasi-non-invasive tool to set up precision medicine strategy for cancer patients. Tremendous efforts have been made to develop the second-generation of “filtration-based” technologies to detect CTCs, revealing a surprising heterogeneity among those cells. Here, we performed the largest and simultaneous analysis of all atypical circulating tumor cells (*aCTCs*) detected with a filtration-based technology, in a cohort of metastatic breast cancer (mBC) patients, and correlated their presence with clinicopathological and survival data.

**Methods:** The PERMED-01 study enrolled patients with mBC refractory to systemic therapy. We prospectively analyzed *aCTCs* present at the time of inclusion in the study, using the Screencell®Cyto device (n=91). Subsets cut-offs were established and evaluated for correlation with clinicopathological data, including progression-free survival (PFS) and overall survival (OS).

**Results:** The median number of *aCTCs* found in mBC was 8.3 per mL of blood. Three subsets of *aCTCs*, absent from controls, were observed in mBC patients: single (*s-aCTCs*), circulating tumor micro-emboli (CTM), and giant-*aCTCs* (*g-aCTCs*). The presence of *g-aCTCs* was associated with shorter PFS and OS in multivariate analyses. For 23 cases, the analysis was completed with advanced immunofluorescence staining and showed that *CTM* and *g-aCTCs* displayed a hybrid phenotype for epithelial and mesenchymal markers.

**Conclusions:** This study highlights the heterogeneity of *aCTCs* in mBC patients both at the cytomorphological and molecular levels when using a Screencell®Cyto device. It reveals the *g-aCTC* subset as a prognostic factor and a potential stratification tool that might help to orientate late-stage mBC patients’ therapeutic care.

## BACKGROUND

Metastases are responsible for more than 90% of cancer-associated mortality and anticipating and understanding their evolution using simple and reliable biomarkers is paramount. Some tumor cells (shed from primary or metastatic tumors) can enter the blood circulation and, in this circumstance, are specifically called “Circulating Tumor Cells” (CTCs). A few of them, fit enough to survive to the drastic conditions encountered in the blood flow (anoikis, sheer stress, anti-tumor immunity…), will eventually spread to other organs and might initiate, after a variable period of dormancy, novel malignant foyers [1].

CTCs sampling is minimally invasive and can be repeated on demand. It is an “*easy*” access to distant tumor lesions. In this line, in the early 2000s, the Cellsearch™ technology (Menarini-Silicon Biosystems, Italy) was FDA-approved to count CTCs. Thanks to this technology, CTCs enumeration at diagnosis is now recognized as an independent predictor of progression-free survival (PFS) and overall survival (OS) in metastatic solid tumors such as breast, colon, and prostate cancers [2, 3]. In metastatic breast cancer (mBC) patients, variation in CTC counts during chemotherapy is predictive for disease progression and survival on a real-time basis, as therapy proceeds [3, 4]. Nonetheless, CTCs prognostic value has a poor added clinical utility in metastatic patients and remains unproven in non-metastatic patients [2, 5]. Second-generation technologies for CTCs detection, based on cell size and deformability, have then emerged.

These new technologies revealed a surprising layer of complexity, showing that CTCs detected with the CellSearch™ represent a small fraction of a more heterogeneous pool of CTCs. Based on cytomorphological characteristics, and until further proper sub-classification is available, any atypical cellular event with malignant morphological features (*ie*. enlarged nuclei, irregular nuclear borders, anisonucleosis, abnormal nucleocytoplasmic ratio) is referred to as an “*atypical Circulating Tumor Cell*” or *aCTC*. Some of the cells found in the blood of cancer patients, but not in healthy donors, can indeed be very atypical, also regarding the cell type inference [6]. The current hypothesis is that the total count of a*CTCs* might dampen their prognostic value because of the intrinsic biological heterogeneity of the population considered [7]. And indeed, fragmented data from small cohorts of patients with solid malignancies have reported the presence of at least three totally different subsets of a*CTCs* [8–10]. They were named after their main cytological presentation: single-atypical CTC (*s-aCTC*), circulating tumor micro-emboli (*CTM*) and giant-atypical CTC (*g-aCTC*).

Here, we decided to study this heterogeneity and to report the prognostic value for each *aCTCs* subset in mBC refractory to systemic therapy. For this, we used the Screencell®Cyto device (ScreenCell™, France), one of the promising and easy-to-use filtration-based technologies.

## MATERIALS AND METHODS

### PERMED-01 sub-study

The PERMED-01 study was a prospective monocentric clinical trial, promoted by and conducted at the Paoli-Calmettes Institute (Marseille, France), and registered as identifier NCT02342158 at the ClinicalTrials.gov platform. More details are available in the **Additional file 1: Materials and Methods**. The present work is an ancillary study of the PERMED-01 assay and is oriented on the analysis of aCTCs subsets isolated from the blood of patients with mBC refractory to at least one line of systemic therapy and with an accessible lesion to biopsy at the time of inclusion [11]. The trial was approved by the French National Agency for Medicine and Health Products Safety, a national ethics committee (CPP Sud-Méditerranée) and our Institutional Review Board. It was conducted in accordance with the Good Clinical Practice guidelines of the International Conference on Harmonization. All patients gave their informed consent for inclusion, biopsy and blood sampling, and molecular analysis. A total of 91 adult female patients, enrolled between January 2015 and December 2016, were sampled for aCTC analysis. For each subject, a blood sample of 5 mL was collected in a Vacutainer® tube containing EDTA K2. The first milliliters of blood were discarded to avoid endothelial cells contamination during the puncture. All samples were shipped at 4°C to the laboratory and processed extemporaneously upon reception, within 4 hours.

### Atypical circulating cells enrichment using ScreenCell®CYTO device

The Screencell®Cyto device is an approved CE-IVD test. Blood samples were processed with this device, as described elsewhere [8, 12]. In brief, 3 mL of peripheral blood were incubated with a red blood cell lysis/fixative buffer for 8 min at room temperature (RT). A low pressure allows the suspension to pass through a metal-rimmed filter, dotted with 7.5+/-0.4 µm diameter pores. The metal-rimmed filter was rinse with Phosphate Buffered Saline (PBS), dislodged from the device, then dried at RT before use with May Grünwald Giemsa (MGG) and/or immunofluorescence staining.

### Atypical circulating cells staining with May Grünwald Giemsa (MGG)

Filters were stained using an MGG kit (Merck Millipore, France) according to the manufacturer’s instructions. Afterwards, filters were air-dried at RT for 5 minutes and stored protected from light until subsequent analysis by light microscopy using a Leica™ microsystem light microscope (10X, 20X and oil-40X objectives) and the NIS-Elements Viewer software (Nikon®). The complete screening of the metal-rimmed filter, which can be performed retrospectively, takes 10 to 15 minutes for an experienced eye.

### Atypical circulating cells subsets according to cytological criteria by MGG

To avoid biases in the comparative analysis of samples, the screening of processed filters was done by two investigators (EM and AL). Every potential *aCTC* was documented and categorized by the trained cytologists, as previously described [8]. Correlation between cytologists was above 92% concordance, and questionable interpretations were selected for discussion until a consensus was reached. In very rare occasions, the event was considered as “uncertain” and thus excluded from the analysis (**Additional file 2: Figure S1**).

Potential *aCTCs* were identified using the following criteria: cell size and shape, irregularity of nuclear borders, enlarged nucleus size, anisonucleosis, dense hyperchromatic nucleus, not totally opaque, nucleocytoplasmic (N/C) ratio [8, 13–15]. Cells without visible cytoplasm were not included in this study. Details about the criteria used for each subset are listed in **Fig. 1a-c** and in **Additional file 3: Table S1**. Briefly, *s-aCTCs* were identified as epithelioid cells with enlarged irregular hyperchromatic nuclei (≈20+/-4 μm) and a high nucleocytoplasmic (N/C) ratio (>0.75). CTM identification criteria were based on the cluster of at least three cells, with nuclei showing signs of anisonucleosis, often with irregularity of nuclear borders and dense hyperchromatic nuclei [12, 14, 16]. The *g-aCTCs* were individualized very large cells (≈50-300µm) identified based on a voluminous cytoplasm (low N/C ratio), which can be round or oblong, and with an enlarged nuclear profile (>20μm in diameter), often multilobular or with separated polymorphic nuclei [17, 18].

**FIGURE 1:**
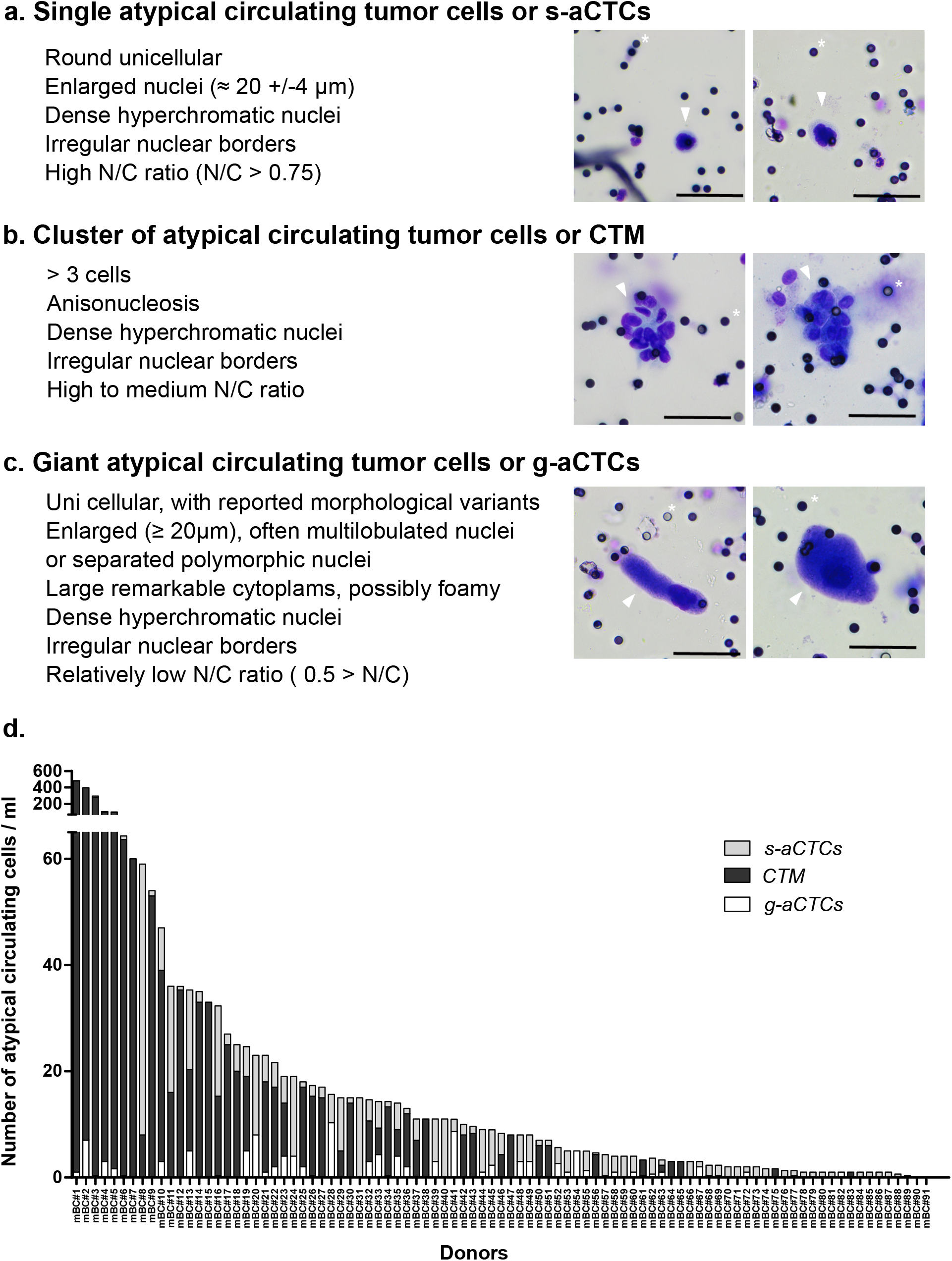
Images of atypical circulating cells and respective subsets distribution in mBC patients. **a-c** May-Grünwald staining of *aCTCs* isolated from the blood of mBC patients using the Screencell®Cyto device. Small black dots are filter’s pores (for clarity some examples are marked with a white asterisk). Scale represents 50µm. Cells of interest are marked with an arrow. The figure shows representative examples of: **a** Single cell or *s-aCTCs*, **b** Cluster of cells or *CTM*, **c** Giant cells or *g-aCTCs*. **d** Distribution of aCTC subsets in the mBC patients (n=91). Number of *s-aCTC, CTM* and g*-aCTC are indicated* per mL of blood.

### Immunofluorescence staining of aCTCs for confocal microscopic analysis

We set up a 6-color immunofluorescence staining simultaneously targeting leukocytes (CD45), epithelial markers (EPCAM and Pan-cytokeratin: pan-KRT), mesenchymal marker (VIM), the stem cell marker LGR5, the efflux pump ABCB1 and SytoxBlue as a DNA labeling dye. Antibodies are detailed in **Additional file 3: Table S2**. All antibodies used in the combination were first validated on cell lines with known positive or negative expression for these markers. Antibodies specificity, signal/noise ratio and antibodies combination are shown in **Additional file 2: Figure 2a-c**.

**FIGURE 2:**
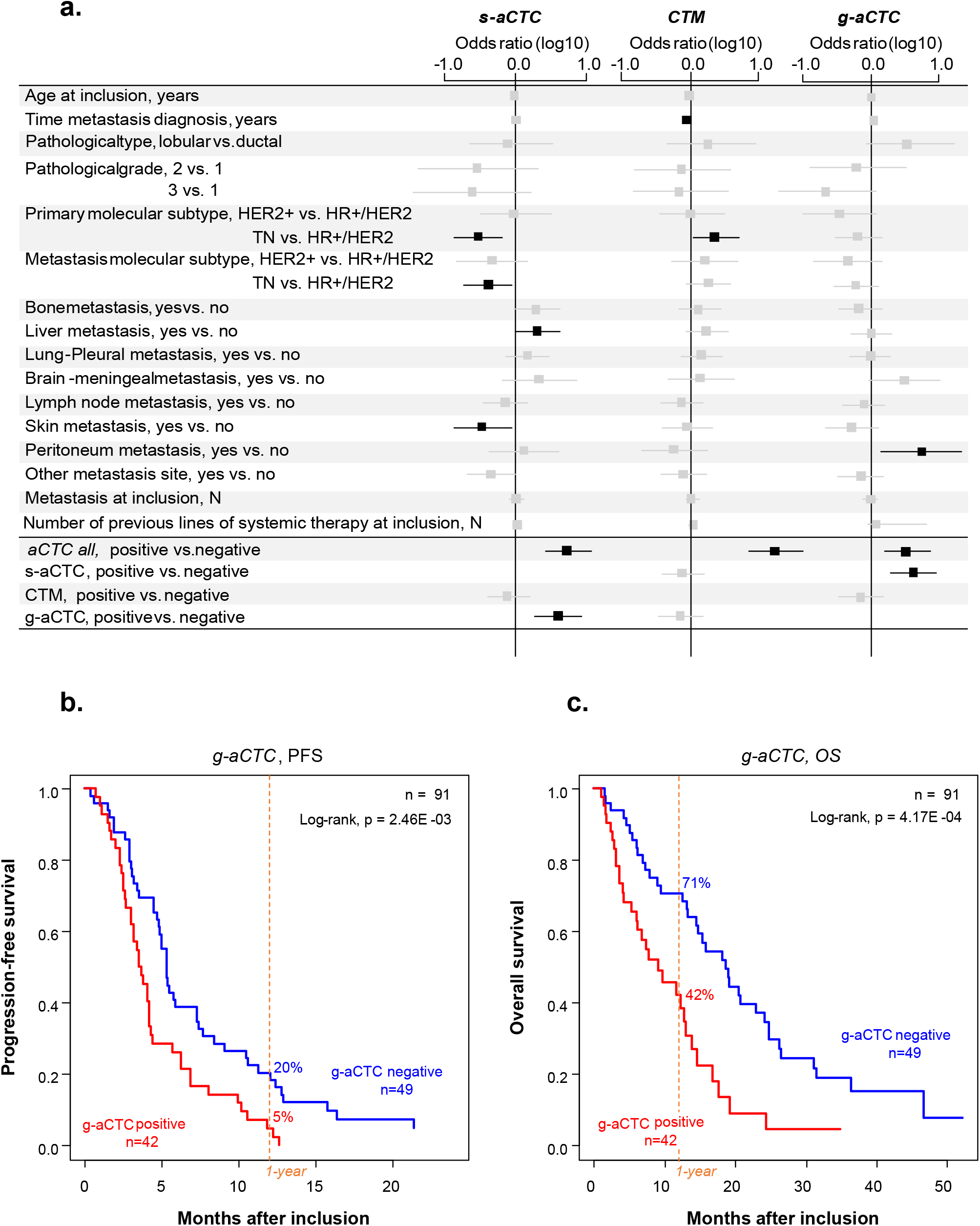
Correlations between aCTC subsets and clinicopathological variables and survival data. **a** Forest plot representation of the correlation between the total number of atypical cells detected, or individual atypical cell subsets (*s-aCTCs, CTM, g-aCTCs*), and clinical data for all 91 patients. Odds ratio are indicated with confidence intervals (horizontal lines). Data statistically significant are in black, data statistically insignificant (crossing 0, vertical line) in grey. TN: triple-negative, HR+: Hormone Receptive-positive, HER2-/+: HER2 negative/positive. The bottom part of the forest plot shows the correlation between the total number of atypical cells and the different subsets. **b-c** Kaplan-Meier PFS (**b**) and OS (**c**) curves of mBC patients according to the presence of *g-aCTCs*. Data are represented as mean ± SEM (n = 3). Differences were considered significant at p < 0.05 (*), p < 0.01 (**), and p < 0.001 (***).

Immunofluorescence staining was done on 23 filters, fixed in paraformaldehyde 4% for 5 minutes at RT, rinsed 3 times in PBS, dried then stored at 4°C until use (stopping point). Filters were then re-hydrated in TBS and permeabilized in TBS-0.2% Triton-X100 for 5 minutes on the day of use. After a quick rinse in water, the filter was incubated in blocking buffer (3% Bovine Serum Albumin, 1% Donkey serum and 1% Goat serum) at RT for 30 minutes to minimize non-specific staining. Primary antibodies were added in blocking buffer and incubated overnight at 4°C. Following three washes with TBS 0.05% Tween20 (TT20), the secondary antibody mixture was added for 1 hour in the dark at RT. After three washes with TT20, the anti-CD45-A488 antibody was incubated for 1 hour in the dark at RT. Finally, after three washes with TT20 and water, the filter was counterstained with Sytox Blue Nucleic Acid Stain for 5 minutes at RT in the dark (Life Technologies). Finally, the filters were mounted with Kaiser Solution (Sigma Aldrich) and dried at RT a few hours. Immunofluorescence was analyzed with a c-Plan-Apochromat 40x/1.3 oil objective on a LSM880 confocal with spectral detection from Zeiss equipped with a 405-laser diode, an Argon-laser and 561- and 633-lasers. Images acquisition and spectra unmixing were done using the Zen Black software.

### Statistical analysis

Correlations between aCTC subsets and clinicopathological variables were established using logistic regression (Logit link function). PFS and OS were calculated from the date of inclusion in PERMED-01 until the date of first progression or death from any cause. Follow-up was measured from the date of inclusion to the date of last news for event-free patients. Survivals were calculated using the Kaplan-Meier method and curves were compared with the log-rank test. Uni- and multivariate prognostic analyses were done using Cox regression analysis (Wald test). The variables submitted to univariate analyses included patients’ age at inclusion, metastasis-to-diagnosis time, pathological type, grade, and molecular subtype of primary tumor, molecular subtype of metastasis biopsied in PERMED-01, nature and number of metastatic sites at inclusion, number of previous lines of chemotherapy at inclusion, and *aCTC* subsets. Multivariate analyses included the variables significant in univariate analysis (p≤0.05). The cell subset discretization was based on the distribution of count cells values of each *aCTC* subset independently, and cut-offs were established statistically using a two-component Gaussian finite Mixture Model (GMM) using maximum likelihood estimation on a per-study basis as previously described [19]. The likelihood ratio (LR) tests were used to assess the prognostic information of one variable provided beyond that of another variable, assuming a χ2 distribution. Changes in the LR values (ΔLR-χ2) measured quantitatively the relative amount of information of one model compared with another. All statistical tests were two-sided at the 5% level of significance. Statistical analysis was done using the survival package (version 2.30) in the R software (version 3.5.2; http://www.cran.r-project.org/).

## RESULTS

### Patients’ population

Ninety-one mBC patients enrolled in the PERMED-01 trial were included in the present study. Patients’ median age at inclusion in PERMED-01 was 55 years (range, 27-79) (**Table 1**). The median time between diagnosis of metastatic relapse and primary cancer was 3 years (range, 0.6-23). Regarding the primary tumor, the most frequent pathological type was ductal (93%), the most frequent pathological grade was 3 (50%), and the molecular subtypes were mainly HR+/HER2- (56%), then triple-negative (TN) (32%), and HER2+ (12%). The molecular subtype of the PERMED-01 metastatic biopsies, available for 87 out of 91 patients, included more TN cases (46%) than HR+/HER2- (41%) and HER2+ (13%). Nineteen patients (22%) showed discordance between the molecular subtypes observed in the metastatic samples and the primary tumor. The median number of metastatic sites at inclusion was 3 (range, 1-8). The most frequent sites were bone (65%), then lymph node (63%) and liver (57%). The median number of lines of systemic treatments received before inclusion was 4 (range, 1-10). Regarding chemotherapy, the median number of previous lines received after inclusion was 3 (range, 1-7), and as expected, 91% of patients had previously received Taxane and/or Anthracycline treatments. The systemic treatments received before and after inclusion are summarized in **Additional file 3: Table S3**. With a median follow-up of 12 months after inclusion (range, 1-52), all but one patient showed disease progression and 68 died. The 1-year PFS was 13% (95%CI 8-22), median PFS was 5 months (range, 1-47), 1-year OS was 58% (95%CI 40-70) and median OS was 14 months (range, 1-52).

**Table 1:**
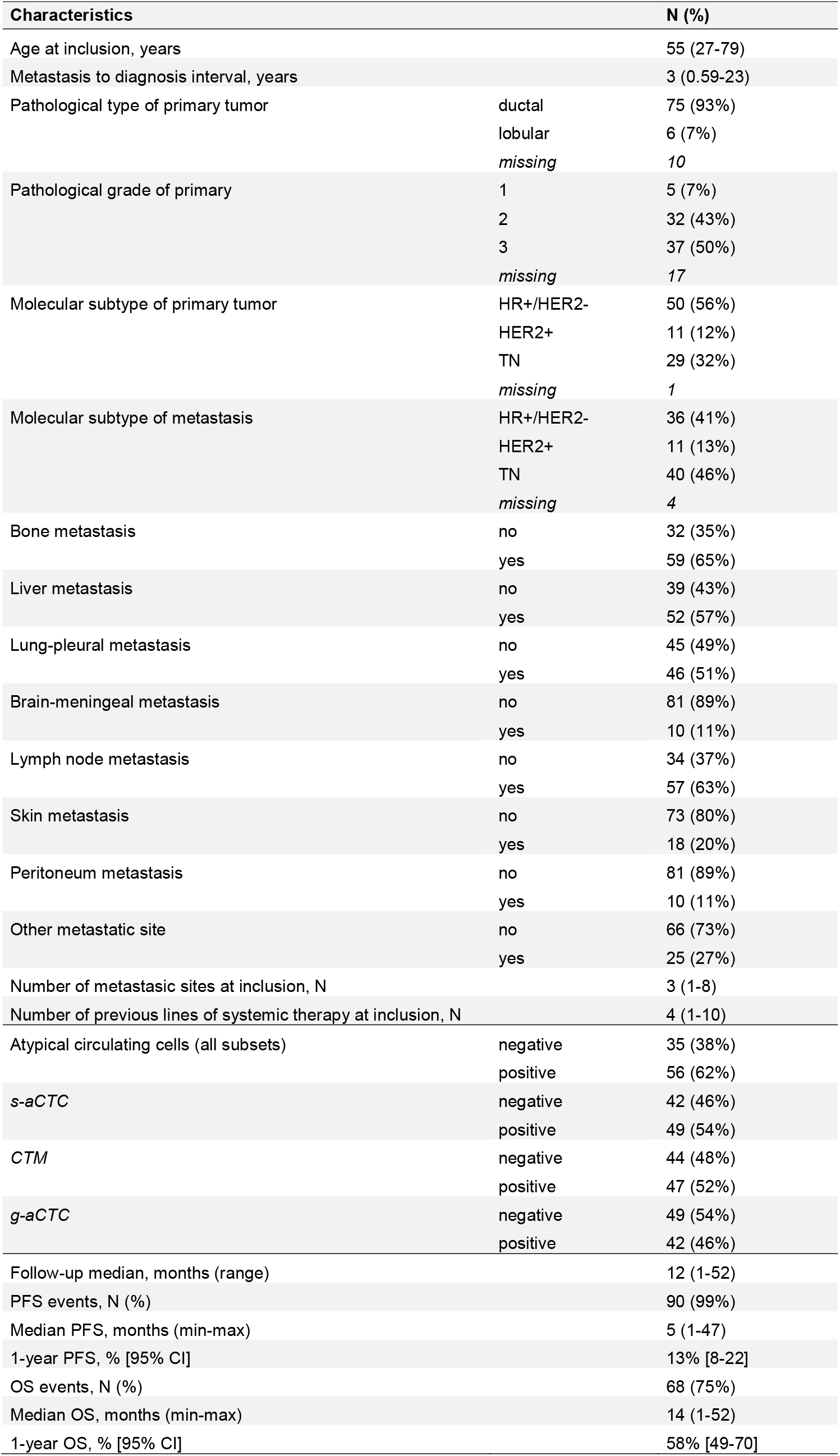
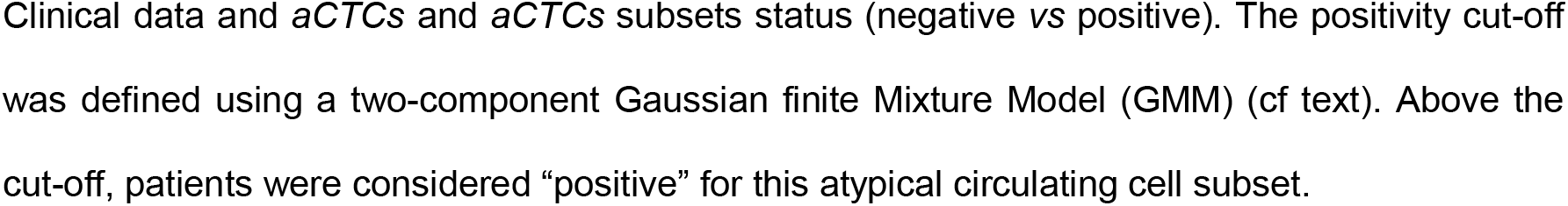
Patient and tumor characteristics.

| Characteristics |  | N (%) |
| --- | --- | --- |
| Age at inclusion, years |  | 55 (27-79) |
| Metastasis to diagnosis interval, years |  | 3 (0.59-23) |
| Pathological type of primary tumor | ductal | 75 (93%) |
|  | lobular | 6 (7%) |
|  | missing | 10 |
| Pathological grade of primary | 1 | 5 (7%) |
|  | 2 | 32 (43%) |
|  | 3 | 37 (50%) |
|  | missing | 17 |
| Molecular subtype of primary tumor | HR+/HER2- | 50 (56%) |
|  | HER2+ | 11 (12%) |
|  | TN | 29 (32%) |
|  | missing | 1 |
| Molecular subtype of metastasis | HR+/HER2- | 36 (41%) |
|  | HER2+ | 11 (13%) |
|  | TN | 40 (46%) |
|  | missing | 4 |
| Bone metastasis | no | 32 (35%) |
|  | yes | 59 (65%) |
| Liver metastasis | no | 39 (43%) |
|  | yes | 52 (57%) |
| Lung-pleural metastasis | no | 45 (49%) |
|  | yes | 46 (51%) |
| Brain-meningeal metastasis | no | 81 (89%) |
|  | yes | 10 (11%) |
| Lymph node metastasis | no | 34 (37%) |
|  | yes | 57 (63%) |
| Skin metastasis | no | 73 (80%) |
|  | yes | 18 (20%) |
| Peritoneum metastasis | no | 81 (89%) |
|  | yes | 10 (11%) |
| Other metastatic site | no | 66 (73%) |
|  | yes | 25 (27%) |
| Number of metastatic sites at inclusion, N |  | 3 (1-8) |
| Number of previous lines of systemic therapy at inclusion, N |  | 4 (1-10) |
| Atypical circulating cells (all subsets) | negative | 35 (38%) |
|  | positive | 56 (62%) |
| s-aCTC | negative | 42 (46%) |
|  | positive | 49 (54%) |
| CTM | negative | 44 (48%) |
|  | positive | 47 (52%) |
| g-aCTC | negative | 49 (54%) |
|  | positive | 42 (46%) |
| Follow-up median, months (range) |  | 12 (1-52) |
| PFS events, N (%) |  | 90 (99%) |
| Median PFS, months (min-max) |  | 5 (1-47) |
| 1-year PFS, % [95% CI] |  | 13% [8-22] |
| OS events, N (%) |  | 68 (75%) |
| Median OS, months (min-max) |  | 14 (1-52) |
| 1-year OS, % [95% CI] |  | 58% [49-70] |

Clinical data and *aCTCs* and *aCTCs* subsets status (negative vs positive). The positivity cut-off was defined using a two-component Gaussian finite Mixture Model (GMM) (cf text). Above the cut-off, patients were considered “positive” for this atypical circulating cell subset.

### Three subsets of aCTCs are found in the blood of mBC patients

Very few data provide simultaneous characterization of the three subsets of *aCTC* previously found in the blood of cancer patients. We wondered whether an identification based on cytomorphological criteria only, as described in [8,12], could be a way, rapid for an experienced eye and independent from markers, to identify *aCTCs* subsets in mBC patients. We thus screened 91 blood samples from advanced mBC for the presence of *aCTCs* after MGG coloration of a ScreenCell®Cyto filter. We observed that three subsets of *aCTCs*, previously described in other solid tumors and on some rare occasions in mBC, were present in the blood of mBC patients. These *aCTCs* present themselves as single cells (*s-aCTCs)* **(Fig. 1a)**, clusters of cells (*CTM)* **(Fig. 1b and Additional file 2: Figure S1a)** or giant cells (*g-aCTCs)* **(Fig. 1c)**.

### The count of aCTC subsets is highly variable in mBC patients

Next, we characterized the frequency and distribution of each subset individually and for each patient (**Fig. 1d, Additional file 3: Table S4)**. All but four patients (96%) had at least one *aCTC* (defined as the sum of *s-aCTCs, CTM*, and *g-aCTCs*) per mL of blood. The median number of *aCTCs* per mL was 8.33 (range, 0-481.6). The median number of *s-aCTCs* per mL was 2 (range, 0-51), with 72 patients with at least one *s-aCTC* per mL (79%). The median number of *CTM* was 1.33 per mL (range, 0-479.6), with 47 patients with at least one *CTM* per mL (52%). The median number of *g-aCTC* per mL was 0 (range, 0-10.3), with 33 patients with at least one *g-aCTC* per mL (36%). The distribution of the different cell subsets was variable between patients: *s-aCTCs* were more frequent than *CTM*, which were more frequent than *g-aCTCs*. When present, the *CTM* and *s-aCTCs* were in larger concentration per mL of blood than were *g-aCTCs*.

Based on the distribution of each *aCTC* subset, we defined objectively a positivity cut-off using a two-component Gaussian finite Mixture Model in the prospect of establishing correlations between all or each *aCTCs* subsets and clinicopathological data. Above the cut-off, patients were considered “positive” for this cell subset. The number of positive patients was 41 (45%) for all *aCTCs* (cut-off: >4.5 cells/mL), 49 (54%) for *s-aCTCs* (cut-off: ≥1.8 cells/mL), 22 (24%) for *CTM* (cut-off: ≥0.9 cells/mL), and 42 (46%) for *g-aCTCs* (cut-off: ≥0.33 cells/mL) **(Additional file 3: Table S4)**.

### Different aCTC subsets correlate with different clinicopathological features

Correlations between *aCTCs* or each individual subset cut-off values and clinicopathological features of patients and tumors (logit function test) are listed in **Additional file 3: Table S5**. Even if these results must be interpreted with caution given the high number of statistical tests performed, interesting correlations were nonetheless observed. A number of *aCTCs* above the cut-off was associated with the presence of liver metastases (p=3.13E-02) and with other *aCTC* subsets above their cut-offs. The three cell subsets showed different correlations (**Fig. 2a)**. The presence of *s-aCTCs* was more frequently associated with the HR+/HER2-subtype of primary cancer (p=1.29E-02) and of metastatic lesion (p=3.22E-02), with the presence of liver metastases (p=4.56E-02), the absence of skin metastases (p=2.86E-02) and the positivity of *g-aCTCs* (p=2.29E-03). The presence of *CTM* was associated with a shorter time between metastatic relapse and diagnosis of primary cancer (p=4.51E-02) and the TN *versus* HR+/HER2-subtype of primary cancer (p=3.39E-02). The presence of *g-aCTCs* was associated with the existence of peritoneal metastases (p=3.75E-02) and the positivity of *s-aCTCs* (p=2.29E-03). None of the *aCTC* subset was associated with patients’ age, number of metastatic sites and number of previous lines of chemotherapy at inclusion, and pathological type and grade of primary tumor.

### The g-aCTCs “positive” status is an independent prognostic factor for PFS and OS

We then asked if all *aCTCs* or individual *aCTC* subsets were associated with patients’ survival. We thus performed uni- and multivariate analyses (Wald test; **Table 2**). In univariate analysis, patients with numbers of *aCTCs* above the positivity cut-off (“positive”) displayed shorter PFS than “negative” patients (HR=1.37, 95%CI 0.89-2.10), but the difference was not significant (p=0.158). Such association was also not significant for *s-aCTCs* and for *CTM*. By contrast, it was significant for the *g-aCTCs* subset, with a HR for PFS event of 1.94 (95%CI 1.24-3.01) in the “positive” patients as compared to the “negative” patients (p=2.98E-03). The 1-year PFS was 5% (95%CI 1-18) in the *g-aCTC* “positive” *versus* 20% (95%CI 12-35) in the “negative” group (p=2.46E-03, log-rank test; **Fig. 2b**), and the respective median PFS were of 3.6 months (range, 3-4.3) and 5.3 months (range, 4.8-7.4). The other clinicopathological features associated with PFS in univariate analysis included the patients’ age at inclusion (p=3.34E-02) and the number of previous lines of chemotherapy at inclusion (p=3.48E-02). In multivariate analysis, the *g-aCTC* status remained significantly associated with shorter PFS (p=2.19E-02), suggesting independent prognostic value.

**Table 2:** Univariate and multivariate analyses of PFS and OS. *#: variables tested as continuous values*

|  |  |  | PFS |  |  |  |  |  | OS |  |  |  |  |  |
| --- | --- | --- | --- | --- | --- | --- | --- | --- | --- | --- | --- | --- | --- | --- |
|  |  |  | Univariate |  |  | Multivariate |  |  | Univariate |  |  | Multivariate |  |  |
|  |  |  | N | HR [95%CI] | p-value | N | HR [95%CI] | p-value | N | HR [95%CI] | p-value | N | HR [95%CI] | p-value |
| Age at inclusion<sup>#</sup>, years |  |  | 91 | 1.02 [1.002-1.04] | **0.0334** | 91 | 1.02 [1.00-1.04] | **0.0378** | 91 | 1.01 [0.99-1.04] | 0.286 |  |  |  |
| Metastasis to diagnosis interval<sup>#</sup>, years |  |  | 91 | 1.02 [0.98-1.06] | 0.34 |  |  |  | 91 | 1.03 [0.99-1.08] | 0.173 |  |  |  |
| Pathological type of primary tumor | lobular vs. ductal |  | 81 | 1.24 [0.53-2.86] | 0.621 |  |  |  | 81 | 1.60 [0.57-4.49] | 0.371 |  |  |  |
| Pathological grade of primary | 2 vs. 1 |  | 74 | 0.74 [0.28-1.97] | 0.453 |  |  |  | 74 | 0.64 [0.19-2.18] | 0.533 |  |  |  |
|  | 3 vs. 1 |  |  | 0.59 [0.23-1.55] |  |  |  |  |  | 0.53 [0.16-1.80] |  |  |  |  |
| Molecular subtype of primary | HER2pos |  | 90 | 1.47 [0.74-2.91] | 0.151 |  |  |  | 90 | 1.50 [0.67-3.39] | 0.262 |  |  |  |
|  | TN |  |  | 1.99 [0.96-4.12] |  |  |  |  |  | 1.98 [0.85-4.64] |  |  |  |  |
| Molecular subtype of metastasis | HER2pos |  | 87 | 0.87 [0.43-1.76] | 0.866 |  |  |  | 87 | 1.26 [0.58-2.73] | 0.694 |  |  |  |
|  | TN |  |  | 1.05 [0.67-1.67] |  |  |  |  |  | 1.25 [0.73-2.14] |  |  |  |  |
| Bone metastasis | yes vs. no |  | 91 | 0.89 [0.57-1.38] | 0.591 |  |  |  | 91 | 1.04 [0.62-1.74] | 0.892 |  |  |  |
| Liver metastasis | yes vs. no |  | 91 | 0.81 [0.52-1.25] | 0.335 |  |  |  | 91 | 1.01 [0.62-1.66] | 0.958 |  |  |  |
| Lung-pleural metastasis | yes vs. no |  | 91 | 1.09 [0.72-1.66] | 0.679 |  |  |  | 91 | 1.06 [0.65-1.71] | 0.817 |  |  |  |
| Brain-meningeal metastasis | yes vs. no |  | 91 | 1.42 [0.73-2.75] | 0.304 |  |  |  | 91 | 3.15 [1.58-6.28] | **0.00115** | 91 | 2.74 [1.37-5.48] | **0.00454** |
| Lymph node metastasis | yes vs. no |  | 91 | 1.06 [0.68-1.65] | 0.784 |  |  |  | 91 | 1.14 [0.68-1.91] | 0.607 |  |  |  |
| Skin metastasis | yes vs. no |  | 91 | 1.02 [0.60-1.74] | 0.942 |  |  |  | 91 | 1.43 [0.81-2.51] | 0.219 |  |  |  |
| Peritoneum metastasis | yes vs. no |  | 91 | 1.20 [0.62-2.32] | 0.597 |  |  |  | 91 | 1.50 [0.71-3.16] | 0.287 |  |  |  |
| Other metastatic site | yes vs. no |  | 91 | 0.95 [0.59-1.53] | 0.845 |  |  |  | 91 | 1.34 [0.80-2.23] | 0.269 |  |  |  |
| Number of metastatic sites at inclusion<sup>#</sup>, N |  |  | 91 | 1.00 [0.86-1.15] | 0.948 |  |  |  | 91 | 1.12 [0.96-1.30] | 0.158 |  |  |  |
| Number of previous lines of systemic therapy at inclusion<sup>#</sup>, N |  |  | 91 | 1.10 [1.01-1.21] | **0.0348** | 91 | 1.06 [0.96-1.16] | 0.23193 | 91 | 1.11 [1.01-1.23] | **0.0396** | 91 | 1.09 [0.98-1.20] | 0.10355 |
| all atypical circulating cells | positive vs. negative |  | 91 | 1.37 [0.89-2.10] | 0.158 |  |  |  | 91 | 1.50 [0.91-2.49] | 0.115 |  |  |  |
| s-aCTC | positive vs. negative |  | 91 | 1.51 [0.99-2.29] | 0.056 |  |  |  | 91 | 1.51 [0.93-2.45] | 0.093 |  |  |  |
| CTM | positive vs. negative |  | 91 | 1.16 [0.77-1.76] | 0.478 |  |  |  | 91 | 1.35 [0.83-2.20] | 0.221 |  |  |  |
| g-aCTC | positive vs. negative |  | 91 | 1.94 [1.25-3.01] | **0.00298** | 91 | 1.87 [1.19-2.95] | **0.00661** | 91 | 2.46 [1.47-4.12] | **0.000584** | 91 | 2.23 [1.31-3.78] | **0.00298** |

Similar results were observed with OS. In univariate analysis, the patients’ status for all *aCTCs, s-aCTCs*, and *CTM* was not significantly associated with OS (**Table 2**). By contrast, it was significant regarding *g-aCTCs* with a HR for death of 2.46 (95%CI 1.47-4.12) in the “positive” *versus* “negative” patients (p=5.84E-04). As shown in **Figure 2c**, the 1-year OS was 71% (95%CI 59-85) in the “*g-aCTC*-negative” group *versus* 42 (95%CI 29-62) in the “*g-aCTC*-positive” group (p=1.06E-04, log-rank test). The respective median OS were 9.2 months (range, 6.1-13.9) and 18.7 months (range, 14.5-24.8). The other variables associated with OS in univariate analysis included the presence of brain/meningeal metastases (p=1.15E-03), and the number of previous lines of systemic therapy at inclusion (p=5.84E-04). Again, in multivariate analysis, the *g-aCTC* status remained significant and associated with shorter OS (p=2.98E-03), suggesting independent prognostic value.

### A hybrid Epithelial-Mesenchymal phenotype is associated with LGR5 and ABCB1 markers co-expression

Because each *aCTC* subset correlated with different clinico-pathological features and had different prognostic values, we wondered whether each subset might display distinct phenotypes. We analyzed *aCTC* subsets more in-depth by looking at their EMT status (EPCAM, Pan-KRT and VIM) and the expression of LGR5 and ABCB1 markers. These markers are often modulated in cells with a mesenchymal-biased profile [20–23], with stemness attributes [24] or resistance to systemic treatments [25]. This list of markers is of course not exhaustive but aims at highlighting potential functional traits observed in progressing diseases, a characteristic of our cohort of patients. The *aCTCs* from the last 23 patients of the cohort were analyzed using advanced multicolor confocal microscopy. This analysis involved a total of 2,152 *aCTCs* (*n=336 s-aCTCs*, n=1,742 *CTM*, and n=74 *g-aCTCs*). The staining observed with the simultaneous combination of these markers, for each *aCTC* subset, is unprecedented (**Fig. 3a** and **Additional file 2: Figure S3**).

**FIGURE 3:**
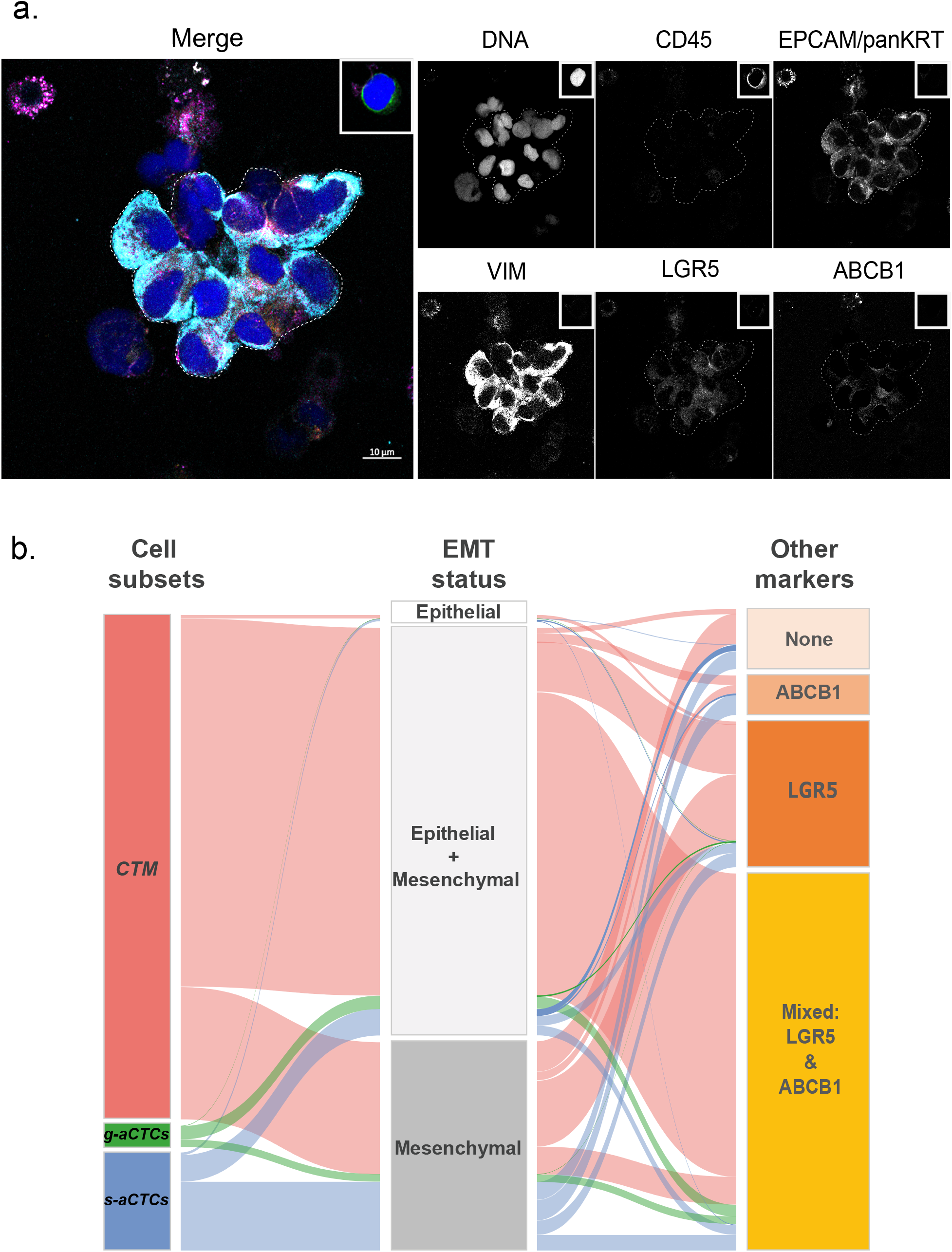
Example of the immunofluorescence staining of *CTM* isolated on ScreenCell® filters. **a** Cells immobilized on the filter were immunostained. Blood cells are detected with the expression of CD45 antigen at the surface of plasma membrane (internal control of the staining shown in the insert top right corner); aCTCs are detected *via* the expression of epithelial markers (EPCAM and pan-KRT) and mesenchymal marker (VIM). LGR5 and ABCB1 expression are also assessed. This image shows a *CTM* with a hybrid EMT status and weak expression of LGR5 and ABCB1. The staining obtained for a leukocyte is represented in the small square of each channel. Scale bar represents 10µm. **b** Alluvial plot representation of the correlations between aCTC subsets and molecular markers. The graph shows the combined expression of EMT, stem-like and efflux pump markers for the three subsets of atypical cells (*CTM, g-aCTCs*, and *s-aCTCs*). The height of the blocks represents the size of the population: in the left column, the pink block represents the amount of *CTM*, the green one of *g-aCTCs*, and blue one of *s-aCTCs*; in the middle column the white block represents cells with epithelial status, light grey with mixed EMT status, and grey with mesenchymal status; in the right hand side column the light pink block the cells negative for both stem-like and efflux pump markers, the light orange block cells expressing the tested efflux pump, the orange block represents the amount of cells expressing stem maker tested, and the yellow block cells expressing both LGR5 and ABCB1 markers. The thickness of a stream represents the number of cells contained in blocks interconnected by the stream.

An epithelial-strict (EPCAM+ and/or KRT+ but VIM-) phenotype was found on very few *aCTCs*, independently of the subset of origin (2.6% of *s-aCTCs*, 0.6% of *CTM*, and 0% of *g-aCTCs*) (**Fig. 3b** and **Additional file 3: Table S6**). The predominant phenotype of *s-aCTCs* was mesenchymal (,.., 70%). The rest of the *s-aCTCs* (27%) displayed a partial EMT, also called hybrid Epithelial-Mesenchymal (E/M) phenotype. The majority of cells observed in *CTM* and *g-aCTCs* displayed a hybrid E/M phenotype (82.5% and 63.6% respectively); few *CTM* and *g-aCTCs* were mesenchymal-only (16.4% and 27.4% respectively). Altogether, these results show that a hybrid E/M phenotype, and to a lesser extend a mesenchymal phenotype, were the most common phenotype observed in *aCTCs* (**Fig. 3b**). We also looked at LGR5 and ABCB1 markers expression. The *s-aCTCs* did not show any preferential pattern for LGR5 or ABCB1 expression (**Additional file 2: Figure S4a**). More than 80% of cells in *CTM* expressed LGR5 and 65% of them were positive for both LGR5 and ABCB1. Finally, 90% of *g-aCTCs* co-expressed LGR5 and ABCB1, independently of their EMT status (**Fig. 3b, Additional file 3: Table S6 and Additional file 2: Figure S4c**). Altogether and even if these data are preliminary, purely descriptive and require additional tests to confirm the functional status of each subset at this stage, this analysis highlights the phenotypic heterogeneity that parallels the cytomorphological heterogeneity of *aCTCs*.

### A hybrid Epithelial-Mesenchymal phenotype is associated with shorter PFS

As shown in numerous studies, cells with a hybrid E/M phenotype have a higher metastatic potential than other cells due to enhanced survival abilities [26]. In an exploratory analysis, we assessed the prognostic value of *aCTCs’* EMT status in the small subgroup of 23 patients. This status was discretized using a 50% positivity cut-off, and considered as positive when more than 50% of *aCTCs* displayed a hybrid E/M phenotype. In univariate analysis (**Additional file 3: Table S7**), the hybrid E/M phenotype was associated with shorter PFS (HR=2.87, 95%CI 1.08-7.68; p=3.53E-02, Wald test). As shown in **Additional file 2: Figure 5a**, the patients without *g-aCTCs* and without *aCTCs* of intermediate EM status (*g-aCTC* 0 + EM 0) displayed 71% 6-months PFS (95%CI 45-100) *versus* 17% (95%CI 3-100) for patients with both positive status (*g-aCTC* 1 + EM 1) and 40% (95%CI 19-85) for patients not matching these classes (*g-aCTC* 0 + EM 1 and *g-aCTC* 1 + EM0) (p=1.12E-02, log-rank test). Such prognostic complementarity was tested using the likelihood ratio (LR) test: the *g-aCTC* status added prognostic information to that provided by the hybrid E/M phenotype of *aCTCs* (ΔLR-χ^2^=4.08, p=4.34E-02; **Additional file 2: Figure 5b**), and conversely, this later added prognostic information to that provided by the *g-aCTC* status alone (ΔLR-χ^2^=2.89, p=8.9E-02). This preliminary result shows that investigating both the molecular phenotype of *aCTCs* and their cytological aspect could improve the prognostic accuracy of *aCTCs*. Given the very small number of patients, this result must be considered as hypothesis-generating only.

## DISCUSSION

The ScreenCell®Cyto device allows the cytomorphological identification of at least three cellular subsets of *aCTCs* in the blood of mBC patients: *s-aCTCs, CTM*, and *g-aCTCs*. Each subset shows correlations with various clinical variables, but only the *g-aCTCs* had an independent prognostic value for survival.

### Reliability

Single-use filter-based systems provide a strong efficiency in isolating *aCTCs*, while most erythrocytes and leukocytes pass through. Compared to other tools, recent data demonstrate that this single-use filter-based device has one of the best sensitivity/specificity balance to isolate *aCTCs* from the blood of patients with solid tumors [9, 27, 28]. Size-based CTCs separation methods, which rely on cell size and deformability, a parameter that is strongly affect by EMT, might however be biased toward enrichment of mesenchymal *aCTCs* [29].

### Comparison

The cytological subsets described here have been reported elsewhere, but studies that simultaneously looked at the three of them in mBC are scarce, with one exception that is more technical than clinical [16]. In this study *(n=20)*, the prevalence of *single, CTM*, and *giant* circulating tumor cells was respectively of 67%, 27%, and 75% in twenty patients compared to 79%, 52%, and 46% in our population (n=91). Major discrepancies can be noted regarding the prevalence of *CTM* and *g-aCTC*, respectively with a higher and lower prevalence in our study.

Concerning the *CTM*, we verified that clusters of more than three cells were not artificially formed during the filtration process (**Additional file 2: Figure S6**). This point of control and the larger size of the patients’ cohort compared to other studies are in favor of a high prevalence of *CTM* in advanced mBC. Although their prevalence is lower than for *s-aCTCs, CTM* have an increased metastatic potential in BC [30, 31]. In line with this, we found that *CTM* have the highest proportion of cells with a hybrid E/M phenotype and expressed the LGR5 marker. This has been observed by others in tumors with high TNM stage [32]. The hybrid E/M phenotype combines the advantages of the mesenchymal phenotype, which confers increased invasiveness and drug resistance to cancer cells, and the epithelial phenotype, endowed with higher proliferation abilities [33]. In mBC, more *CTM* release and a dynamic shift toward a hybrid E/M phenotype during treatment were correlated with treatment failure and disease progression [34], which is coherent with our results. We were however not able to correlate *CTM* presence to shorter patients’ PFS or OS. Our cohort having received multiple prior systemic therapies, one can hypothesize that the predictive value is less powerful in terminal disease. Analyses of variations between time points might be more informative [35, 36]. Nonetheless, the high prevalence of *CTM* and hybrid E/M phenotype are important and new observations in patients with advanced mBC not responding to treatment, suggesting that persistent *CTM* could be an important prognostic marker during treatment.

Explanations regarding the discrepancy in *g-aCTC* prevalence could be multiple. It is indeed acknowledged that the prevalence of these giant cells in the blood of patients with solid tumor is around 40% [37–39]. This might be due to the volume of blood tested (7.5 vs 3 mL in our case), which double the chance to detect these cells.

Most intriguing is the nature of these giant cells, which have been identified in patients with cancers but not in healthy donors (data not shown and [17]). Different names were however given to them: circulating hybrid E/M cells, giant epithelioid cells, fusion-derived epithelial cancer cells, tumor-macrophage fusion cells (TMFs), macrophage-tumor cell fusion cells (MTFs), or cancer-associated macrophage-like (CAMLs) cells [16, 17, 37, 39–42]. This shows that the origin of *g*-*aCTC* subset is still unclear. Some studies attribute their formation to cellular fusion between tumor cells and other tumor cells (homotypic) or with non-tumorous cells (heterotypic). The fusion’s product supposedly combined the attributes of both fused cells, and results in a cancer cell with a more aggressive phenotype, responsible for shorter survival, increased metastatic and chemoresistance capabilities. Similarly, artificial fusion of tumor cells with macrophages shows enhanced migratory, invasive and metastatic phenotypes *in vivo* [37, 39, 43]. It is interesting to note that if *g-aCTCs* are the result of cells fusion, their potential polyploidy could enhance their chances to adapt to environmental changes, which would participate in the increased tumorigenic potential compared to other subsets of *aCTCs*. Looking at *g-aCTCs* polyploidy in the future might be an interesting marker for cell plasticity.

Other studies, not clearly hypothesizing on the cellular fusion events, have identified these large cells as cells related to the macrophage lineage, describing them in a broader way as giant macrophages (CAMLs or MTF or MFT) that contain phagocytosed epithelial debris [6, 16, 17, 38, 40, 41]. The in-depth study of these cells, at the cellular level, showed that some of the giant cells express CD14 or CD11c, two well recognized markers of the macrophage/myeloid lineage. Both hypotheses (fusion or myeloid origin) require further investigations to clarify the inference of these giant cells.

This would help decipher the role of these giant cells in cancer evolution and potentially refine their role as biomarkers, including in early disease [6]. In this line, a seminal study revealed that the type of treatment received (hormone-treated versus chemotherapy) affects the quantity of CAMLs detected, chemotherapy being associated with increased release of CAMLs into the circulation [17]. The authors suggest that CAMLs may provide a sensitive representation of phagocytosis of cellular debris caused by chemotherapy at the tumor site. In our population, treatments, including chemotherapy, were inefficient. It is thus plausible that CAMLs or *g-aCTCs*, if we assume the overlap between these two populations, are less frequently released because of cancer cells resistance to treatment at metastatic sites. ABCB1 efflux pump is involved in resistance to a number of anticancer agents used for mBC treatment, including anthracyclines, vinca alkaloids, and taxanes, among others [25]. Here, we showed its high prevalence on g-*aCTC* and *CTM* subsets. If ABCB1 expression is indeed related to resistance to chemotherapeutic agents in future functional validation, the combined detection of *aCTCs* and ABCB1 marker might be extremely valuable to estimate the efficiency of a given treatment as the therapy proceeds. However, the molecular part of our study in 23 patients is still very preliminary and warrants future validation in larger series. For now, it should be regarded only as hypothesis-generating.

### Limitation

The major limitation of our study was to rely on cytomorphological criteria to detect *a-CTCs, a*lthough based on stringent criteria. This was to be amenable with clinical practice.

The small number of samples compared to the large range of prior systemic therapies (ranging from 1 to 8) and the heterogeneity in term of molecular subtypes, which likely explains the non-significant prognostic value of tumor grade and molecular subtypes of our cohort, are also obvious weakness. However, although not exhaustive in term of tested variables, our prognostic analysis included the best validated prognostic factors [44], such as relapse-free interval, site of metastases, number of metastatic sites and molecular subtypes. Ideally, similar analyses should be reapplied to larger and more homogeneous series of patients.

Finally, the high sensitivity of the ScreenCell®Cyto device might result in the trapping of non-malignant cells on the filters [45]. As such, endothelial cells, although not supposedly found circulating in the blood, have been identified in rare occasions in cluster, notably in cancer patients [46, 47]. Against all expectations, some of these endothelial clusters displayed cytological traits consistent with malignancy, such as atypical nuclei, prominent nucleoli, and high nuclear-to-cytoplasmic ratio. However, in mBC patients, the analysis of their cell-type inference revealed an epithelial-derived tumor cell origin [31].

### Benefit and practical implication

Our work presents the largest cohort of mBC patients analyzed so far using the ScreenCell®Cyto system. It comforts the validity and utility of this easy, relatively inexpensive and extremely convenient system to study *aCTCs*. Still, it required the expertise of a cytologist for analysis. In this prospect, the development of an automatized detection of *aCTC* subsets is ongoing (Project Medicen, part of the NCT03797053 trial). Validation has already been achieved with samples from melanoma patients, showing a sensitivity of automated counting compared to conventional reading of 97% (11th International Symposium on Minimal Residual Cancer, May 2018).

## Conclusion

*In fine*, these results highlight i) the heterogeneity of *aCTC*s that can be observed in cancer patients, which might include non-malignant cells potentially “coming from the tumor”, and ii) the importance for increasing our knowledge of these subsets to discover new mechanisms involved in oncogenesis. Furthermore, using a *real-life* cohort of advanced mBC patients, we highlighted the *g-aCTCs* subset as an independent prognostic factor in mBC patients, both regarding PFS and OS. This prognostic value might further be improved using the EMT status of this subset. The correlation of *aCTCs* subsets, and notably with *g-aCTC*, with clinical and molecular data is new. Our study suggests at least two clinical added values: the presence of *g-aCTCs* might i) help oncologists to identify which patients, after several lines of systemic therapy, might benefit from best supportive care alone, and ii) be used as a tool for better prognostic stratification in early clinical trials testing novel therapeutics, which frequently enroll late-stage, already heavily-treated and poorly responding patients.

## Data Availability

Data will be available on request

## Abbreviations

*aCTCs*: atypical Circulating Tumor Cells
CAMLs: Cancer-Associated Macrophage-Like cells
CTCs: Circulating Tumor Cells
CTM: Circulating tumor micro-emboli
E/M status: Epithelial to Mesenchymal status
EMT: Epithelial to Mesenchymal Transition
*g-aCTC*: giant-atypical Circulating Tumor Cells
GMM: Gaussian finite Mixture Model
LR: Likelihood Ratio
mBC: metastatic breast cancer
MGG: May Grünwald Giemsa
N/C: nucleocytoplasmic
OS: Overall survival
PBS: Phosphate Buffered Saline
PFS: Progression free survival
RT: room temperature
s-aCTC: single-atypical Circulating Tumor Cells
TAM: Tumor Associated Macrophage
TN: Triple Negative

## Declarations

### Ethics approval statement and consent to participate

The present ancillary study used samples from PERMED-01 prospective assay registered as ClinicalTrials.gov identifier NCT02342158, approved by the French National Agency for Medicines and Health Products Safety, a national ethics committee (CPP Sud-Méditerranée), and Paoli-Calmettes Institute Institutional Review Board (Comité d’Orientation Stratégique, COS). PERMED-01 protocol is accessed on https://clinicaltrials.gov/ct2/show/NCT02342158. All patients gave their informed consent for inclusion, biopsy and blood sampling, and molecular analysis, and patient privacy was protected.

### Consent for publication

Not applicable

### Availability of data and materials

The datasets supporting the conclusions of this article are included within the article (and its additional files). The datasets generated during and/or analyzed during the current study are available from the corresponding author on reasonable request. All patients’ data were anonymized and will be kept as such.

### Author contributions and notes

FB, DB and EM designed the study and obtained the grants to perform the study. AL, LB, SG, AA, MLL, OC, ED, CA and EM performed experiments. PF, AG, QD and FB did the biocomputational analyses. AN, JP, AGo, FB, DB provided patients samples and clinical rationale. FB and EM designed the figures. AL, CA and EM wrote the manuscript. All authors proofread the manuscript. ^#,§^ co-authorship: ^#^ AL and LB equally contributed to this work and should be considered as co-first authors; ^§^ CA, FB and EM equally supervised this work and should be considered as co-last authors

## Acknowledgements

We would like to thank all the donors who have willingly contributed to the study (PERMED-01 Cohort patients, donors without malignant diseases); the Direction de la Recherche Clinique, for the management of the PERMED-01 cohort; D. Isnardon and M. Rodrigues from the Microscopy and Scientific Imaging platform.

## ADDITIONAL FILES

*Additional file 1: Materials and Methods*

### Study objectives and design

PERMED-01 was a prospective unicentric clinical trial promoted by and conducted at the Paoli-Calmettes Institute (Marseille, France). Its primary objective was to evaluate the number of patients with locally advanced or metastatic cancer for whom identification of actionable genetic alterations (AGAs) in tumor samples using t-NGS and aCGH could lead to the delivery of a “matched therapy”. Secondary objectives included the analysis of CTCs (for breast and digestive cancers), but also the assessment of feasibility, toxicity, and incidence on the clinical outcome of prospective molecular screening, the description of molecular alterations of advanced solid cancers and their relationship with the clinicopathological characteristics, their comparison with molecular alterations of the paired primary tumor if available, pan-genomic molecular analysis of metastatic samples with full exome sequencing and transcriptome analysis, analysis of circulating tumor DNA, and development of preclinical models for prediction/analysis of tumor response/resistance (xenografts, short-term culture, and organoids for breast cancer). Inclusion criteria were age ≥18 years, pathological diagnosis of solid cancer, locally advanced or metastatic stage progressive during at least one line of prior therapy and with an accessible lesion for biopsy, Eastern Cooperative Oncology Group (ECOG) Performance Status ≤2, affiliation to Social Insurance, and signed informed patient’s consent for participation. Exclusion criteria were symptomatic or progressive leptomeningeal or brain metastases, bone or brain metastasis as sole metastatic site, pregnancy or breastfeeding, and person in an emergency situation or subject to a measure of legal protection or unable to express consent. The study, registered as ClinicalTrials.gov identifier NCT02342158, was approved by the French National Agency for Medicines and Health Products Safety, a national ethics committee (CPP Sud-Méditerranée), and our Institutional Review Board (Comité d’Orientation Stratégique, COS). It was conducted in accordance to the Good Clinical Practice guidelines of the International Conference on Harmonization. All patients gave their informed consent for inclusion, biopsy, and genomic analysis. Once enrolled, a new tumor biopsy or resection was proposed to the patient.

### Statistical analysis: number of inclusions

In order to have a sufficient number of patients with different cancers and with an identifiable AGA, we wanted to evaluate 300 patients enrolled over three years. Previous studies had reported a 35% technical failure rate. Thus, we initially planned to include 460 patients. On November 2017 after three years of inclusion and inclusion of the 460^th^ patient, and in order to increase the numbers in certain patients’ subpopulations for certain secondary objectives, we asked the French regulatory authorities to amend the protocol to allow enrollment of 100 additional patients over 1 year. The protocol was amended and the trial was reopened to inclusions on September 2018. On September 2019, after one year of inclusion and enrollment of 90 additional patients, the trial was closed. Thus, a total of 550 patients had been enrolled between November 2014 and September 2019, over less than 4 years of inclusion (November 2014 to November 2017, then September 2018 to September 2019).

In the present ancillary study on analysis of CTCs in patients with metastatic breast cancer, a total of 91 adult female patients had been enrolled after disease progression in our trial between January 2015 and December 2016 and were sampled for aCTC analysis. In addition, seven healthy adults lacking without any known pathology were recruited as controls.

### Cell lines

Cell lines were purchased from the ATCC® collection and cultured at 37°C and 5% CO_2_ in recommended media. We used Hs 578T (Hs 578T ATCC ® HTB-126™), SK-BR-3 (ATCC® HTB-30™), SUM-190PT (BioIVT™) and MDA-MB-231 (ATCC® CRM-HTB-26™) breast cancer cell lines, Caco-2 (ATCC® HTB-37™) colon cancer cell line, and K-562 (ATCC® CCL-243™) myeloid cell line. Cell lines were regularly checked for mycoplasma contamination during their growth and discarded in case of positive results.

### Setting up of 6-color immunofluorescence using cell lines

Cell lines seeded on glass coverslips were fixed with 4% paraformaldehyde (Sigma Aldrich) for 5 minutes and permeabilized 5 minutes in TBS containing 0.2% Triton X-100 (Sigma Aldrich). Each antibody used for the multicolor immunofluorescence (**Additional file 3: Table S2**), was independently tested on positive and negative cells to establish the correct dilution and distinguish background from positive staining. EPCAM and pan-KRT antibodies, which are detected in the same channel in the multicolor combination, were also tested together. Multicolor immunofluorescence was conceived with the help of the application note from Zeiss (A ten-color spectral imaging strategy to reveal localization of gut immune cell subsets 2018 and [48]). The combination of secondary antibody fluorophores was chosen to allow spectral distinction (DL405, Sytox Blue, A488, DL549, A647, A680). All secondary antibodies were cross-adsorbed antibodies to minimize background and off-target signals. The correct separation of signals in the combination of primary and secondary antibody was confirmed with control staining on cell lines. The acquisition settings were maintained constant throughout the study and the apparatus performances are ensured by quality control of the imaging platform. The combination of antibodies is shown in **Additional file 3: Table S2**.

### Evidences suggesting that the Screencell®Cyto device do not generate CTM artificially

To address potential artificial formation of clusters, 1,500 “single” cells from a cell line (SK-BR-3 or MDA-MB-231 cells, trypsinized, filtered with 70µm cell strainer and counted) were spiked into 3 ml of blood. This corresponds to the highest concentration of CTCs detected in our cohort. The blood was process within 4 hours with the ScreenCell®Cyto module and the filter was stained by May Grundwald Giemsa.

We then counted the number of clusters observed and documented their size. The result of 3 independent experiments is summarized in **Additional file 2: Figure S6**. Even though CTM can occasionally be observed after the filtration, the vast majority (> 92%) of cells are not observed within CTM.

*CTM* number and size distributions were compared to the data from 3 of our patients, displaying high number of CTM (#318 cells, #1443 cells and #1191 cells in total).

Most interestingly, in patients, the size of CTM was significantly bigger than CTM observed from spiked cells. Additionally, single CTCs (cluster’s size <2 cells) were in minority compared to CTM, opposite to the situation observe in the test experiments. Altogether, this dampens the possibility that the filtration technic artificially induces the formation of cell clusters in general, and especially CTM above 3 cells, on the filter.

In addition, an indirect observation made from patients shows that high number of CTCs is not necessarily associated with the presence of CTM. For example, patient mBC#8 has a large total number of CTCs (60 CTCs/ml), similar to mBC#7 and m#BC9, but does not have many clusters compared to the other two patients. This precludes from artificial formation of CTM in samples with high number of CTCs.

*Additional file 2: Figures*

**Figure S1: Pictures of atypical circulating cells observed in the blood of patients with mBC**.

Cytological staining with May-Grünwald solution of cells isolated from blood samples on a filter. Small black dots are filter pores (for clarity some examples are marked with a white asterisk). Scale represents 50µm. Cells of interest are marked with an arrow. The figure shows representative examples of: **a** Clusters of epithelioid cells of *CTM*, **b** “Uncertain” specimens, including cells with no clearly visible cytoplasm or without morphological malignant features. We observed samples with high frequency of dense and hyperchromatic nuclei (equal or greater than 20 µm) with irregular nuclear borders, but with no visible cytoplasm. These were naked nuclei or residual apoptotic bodies that we referred to as “uncertain” specimens, because the cellular origin cannot be addressed. These events were not analyzed in the present study. Complementary analysis is required to understand their origin: whether the pressure applied during the filtration blows apart the cytoplasm from the nucleus of fragile cells, or the cell is undergoing apoptosis in the bloodstream thereby displaying a retracted cytoplasm. Of note, they were not observed in control samples, and we cannot rule out a potential role in disease progression at this stage. **c** Round naked nuclei or residual apoptotic bodies. Scale represents 50µm. We also occasionally observed in patients and control samples small (<20 µm) round naked nuclei or residual apoptotic bodies that were also not analyzed. Scale represents 50µm.

**Figure S2: Antibody validations**

**a. Antibodies specificity**

Each antibody was tested on positive and negative control cells by immunofluorescence to assess their specificity and the signal/noise ratio.

Pan-Cytokeratin showed a cytoplasmic staining in MDA-MB-231 (and SK-BR-3 not shown) cells. It was more or less filamentous depending on expression levels and cell spreading. Hs 578t cells were negative for cytokeratin staining.

VIM antibody stained the cytoplasm of MDA-MB-231 cells but not SK-BR-3.

Anti-EPCAM antibody stained the plasma membrane of SK-BR-3 cells but not MDA-MB-231. CD45 stained the plasma membrane of hematopoietic malignant cells, K-562, but not epithelial cells, MDA-MB-231.

ABCB1 antibody was positive on Caco2 cells, with a predominant localization at the membrane (https://www.proteinatlas.org/ENSG00000085563-ABCB1) and negative on MDA-MB-231 cells. LGR5 was detected in a fraction of SUM190 cells, but not on MDA-MB-231 cells. LGR5 shows a dotted intracellular staining (Golgi apparatus and nucleoplasm) in agreement with data validated by the human protein atlas website (https://www.proteinatlas.org/ENSG00000139292-LGR5). LGR5 is not detected in MDA-MB231 cells.

Of note, and as expected, markers staining’s in cell lines were always stronger than what we could observe in the majority of cells isolated from patients in our study.

**b. Signal/noise ratio - Background**

MDA-MB-231 cells, which are CD45-breast cancer cell line, grown in suspension (ultra-low adhesion plates) for two days were mixed with blood and filtered on a Screencell®Cyto device. Cells on the filter were immunostained as described in the material and methods to assess the immunolocalization of Pan-KRT and VIM in this condition. It showed that, despite a high expression of the two proteins, the filamentous aspect of Pan-KRT and VIM is less clear in cells in suspension compared to the same cells grown in 2D (**Additional file 2: Figure S2a**).

It also highlights that, from the same cellular suspension, background intracellular CD45 staining can be detected in the cytoplasm of some epithelial cells. In this case, the CD45 labelling is diffuse and faint compared to the plasma membrane labelling observed on a CD45+ leukocyte. On the first line of images, we also added a staining with an anti-CD45 on hematopoietic malignant cells (K-562) to show a true signal against what is a background.

This background “problem” can come from three causes.

First, nonspecific background binding of antibodies (primary and secondary) can give a background signal. To limit this aspect, the saturation step included BSA, donkey and goat serum, and antibodies were titrated to be used at the right concentration for optimal signal to noise ratio. We cannot however control the state of each cells and quality of the filter that also affects the quality of the signal.

The second cause is cellular auto-fluorescence that varies between cells. Auto-fluorescence is stronger and easier to detect on cells with bigger size and with large cytoplasm (in our case in g-*aCTCs* compared to leukocytes). The background is thus different depending on the cell type, cell size and cell shape.

Third, auto-fluorescence is expected to vary with the metabolic activity of cells, and is enhanced in cells copping with severe stresses (auto-fluorescence is due to an increase in production of flavoproteins, which are involved in ROS detoxification). This explain why, within a sample, auto-fluorescence might differ from one cell to another [49]. This is rather intuitive to expect variation in auto-fluorescence of *aCTC* considering the stressful condition they are facing in the circulation.

**c. Antibodies combination validation**

Antibodies targeting different molecules corresponding to a given phenotype, here the epithelial phenotype, were simultaneously used and revealed in the same fluorescent canal.

EPCAM and Pan-KRT antibodies were tested alone or mixed together to check that the combination of both antibodies allows them to conserve their specificity and to enhance the sensibility of detection of the desired phenotype. Scale bars: 10µm.

The combination of all antibodies used on the filters was tested on cell lines to ensure its suitability.

Cells were stained for CD45, Pan-KRT and VIM. Scale bars: 10µm.

**Figure S3: Examples of the immunofluorescence staining of atypical cells isolated on ScreenCell® filters**.

The blood of mBC patients were filtered and cells on the filter were immunostained. Blood cells are detected with the expression of CD45 antigen at the plasma membrane (empty arrowheads), whereas *aCTCs* (marked with dotted lines) are detected *via* the expression of epithelial markers (EPCAM and pan-KRT) and mesenchymal marker (VIM). Stem cell marker (LGR5) and drug resistance marker (ABCB1) expression are also assessed. Representative images of **a**. *s-aCTCs:* CD45-, EPCAM/Pan-KRT-, VIM+, LGR5+, ABCB1+, and CD45-, EPCAM/Pan-KRT-, VIM+, Lgr5+, ABCB1-, **b**. *CTM:* CD45-, EPCAM/Pan-KRT+, VIM+, LGR5+, ABCB1+, and **c**. *g-aCTCs*: CD45 diffuse, EPCAM/Pan-KRT+, VIM+, LGR5+, ABCB1+, and CD45-, EPCAM/Pan-KRT+, VIM+, LGR5+, ABCB1-, are shown and indicated with dotted lines) Empty arrow heads point blood cells (CD45 positive staining the cell surface). Scale bar represents 10µm.

**FigureS4: Alluvial plot representation of the combined expression of EMT, stem and drug resistance markers on aCTCs subsets individually (*CTM, g-aCTCs*, and *s-aCTCs*)**.

The same data as in figure 3 are represented for each cell subset separately. Alluvial plot representation of the correlations between *aCTC* subsets and molecular markers. The graph shows for the three subsets of atypical cells separately (for *s-aCTCs* in **a**, for *CTM* in **b** and for *g-aCTCs* in **c**) the combined expression of EMT, LGR and ABCB1 markers. The height of the blocks represents the size of the population. The thickness of a stream represents the number of cells contained in blocks interconnected by the stream. EMT status: Ep=epithelial, Hybrid=epithelial + mesenchymal, Mes=mesenchymal; Other markers: ABCB1, LGR5, Mixed= ABCB1 + LGR5.

**Figure S5: Kaplan-Meier curves of PFS of patients based on the positivity of *g-aCTCs* and intermediate EM status**.

**a** Survivals were calculated using the Kaplan-Meier method and were compared with the log-rank test to evaluate the prognostic value of combined *g-aCTC* and intermediate EM statuses. Presence of *g-aCTC* is marked as 1 vs absence as 0. Presence of an intermediate EM status in all aCTCs’ subsets is represented as 1 value, absence as 0. **b** Prognostic complementarity: the values are given for prognostic information of each variable colored in grey (Mixed EM and *g-aCTC*) on its own (LR-χ2) and when added to the other variable colored in blue (ΔLR-χ2). * indicates p≤0.05 and indicates trend for significance with p≤0.10.

**Figure S6: Clusters verification**

To address potential artificial formation of clusters during the filtration process 1,500 “single” cells (which corresponds to the highest concentration of *a-CTCs* observed in patient) from a cell line SK-BR-3 or MDA-MD-231 (trypsinized and counted) were mixed into 3 ml of blood. The blood was then process as usually within 4 hours with the ScreenCell®Cyto module and stained with May Grundwald Giemsa to count clusters. The results are compared with cluster occurrence in patient samples containing an equivalent number of *a-CTCs* (318, 1,191 and 1,443 *a-CTCs* in 3mls of blood).

*Additional file 3: Tables*

**Table S1: Summary of the cytomorphological criteria used to define each *aCTC* subset**.

**Table S2: Antibodies used for immunofluorescence**.

**Table S3: Summary of systemic therapies received before and after inclusion**

**Table S4: Distribution of aCTC subsets in the population of mBC patients**.

**Table S5: Correlations between *aCTCs* and *aCTC* subsets and clinicopathological variables**.

**Table S6: Molecular markers expressed by *aCTCs***.

**Table S7: Univariate and multivariate analysis for PFS according to molecular and cytological profiles**.

