## Additional_File2-Figures for "Identification of atypical circulating tumor cells with prognostic value in metastatic breast cancer patients"

### Supplementary Figure 1

**a.** Cluster of epithelioid cells or *CTM*

**b.** "Uncertain" specimens

**c.** Round naked nuclei or residual apoptotic bodies

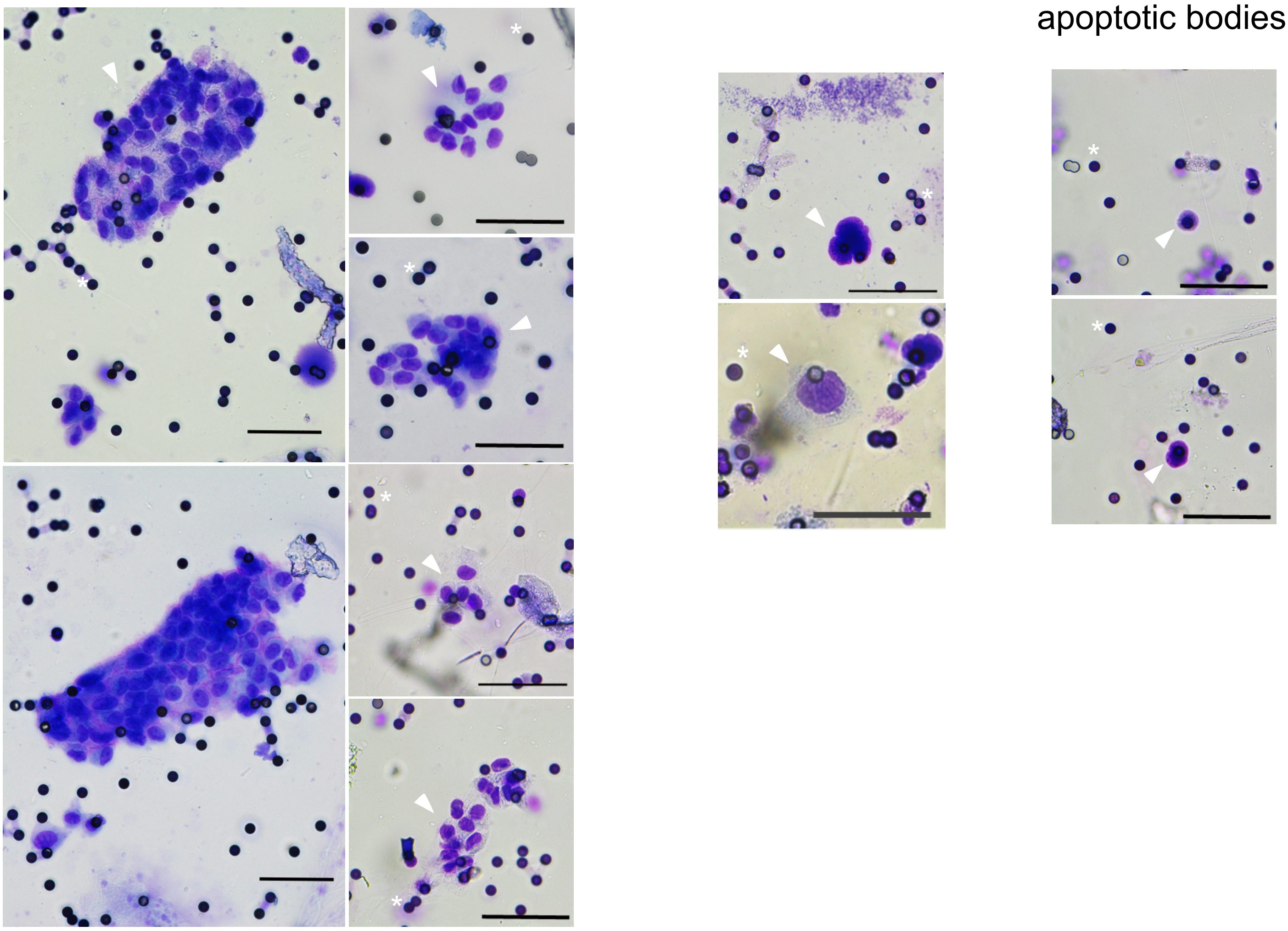

**Figure S1: Pictures of atypical circulating cells observed in the blood of patients with mBC.**

Cytological staining with May-Grünwald solution of cells isolated from blood samples on a filter. Small black dots are filter pores (for clarity some examples are marked with a white asterisk). Scale represents 50µm. Cells of interest are marked with an arrow. The figure shows representative examples of: **a** Clusters of epithelioid cells of *CTM*, **b** “Uncertain” specimens, including cells with no clearly visible cytoplasm or without morphological malignant features. We observed samples with high frequency of dense and hyperchromatic nuclei (equal or greater than 20 µm) with irregular nuclear borders, but with no visible cytoplasm. These were naked nuclei or residual apoptotic bodies that we referred to as “uncertain” specimens, because the cellular origin cannot be addressed. These events were not analyzed in the present study. Complementary analysis is required to understand their origin: whether the pressure applied during the filtration blows apart the cytoplasm from the nucleus of fragile cells, or the cell is undergoing apoptosis in the bloodstream thereby displaying a retracted cytoplasm. Of note, they were not observed in control samples, and we cannot rule out a potential role in disease progression at this stage. **c** Round naked nuclei or residual apoptotic bodies. Scale represents 50µm. We also occasionally observed in patients and control samples small (<20 µm) round naked nuclei or residual apoptotic bodies that were also not analyzed. Scale represents 50µm.

#### Supplementary Figure 2a

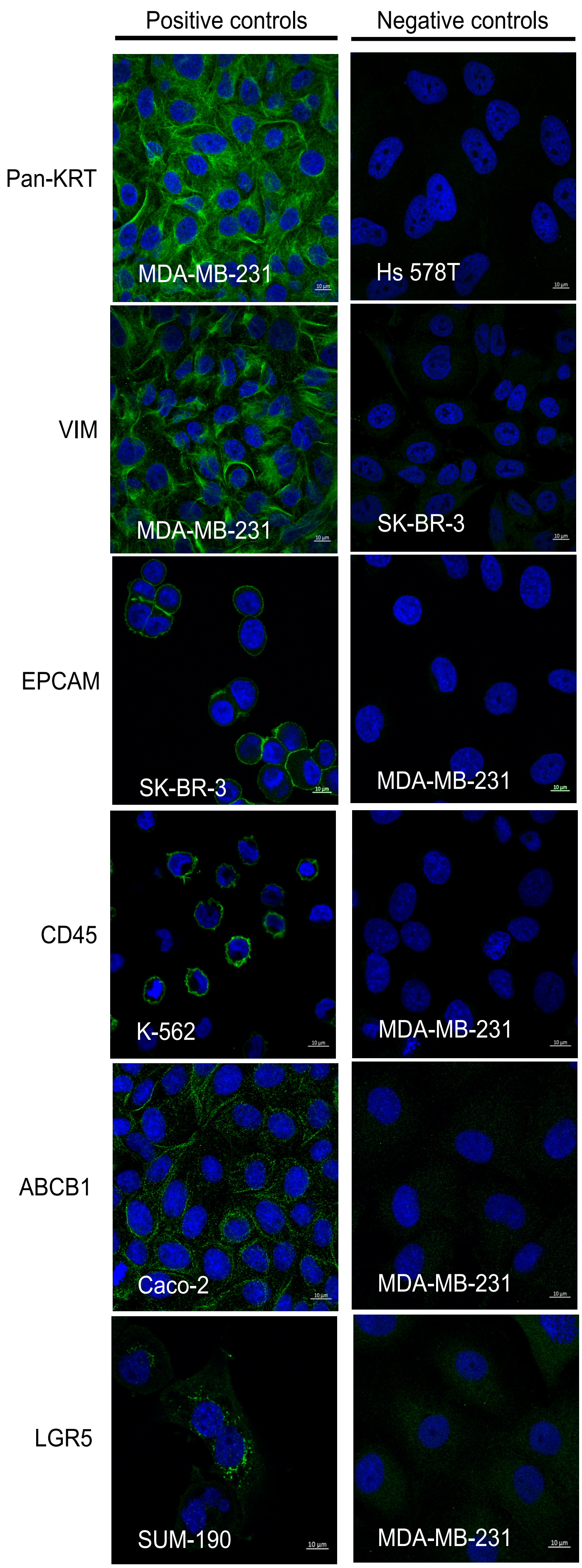

**Supplementary Figure 2b**

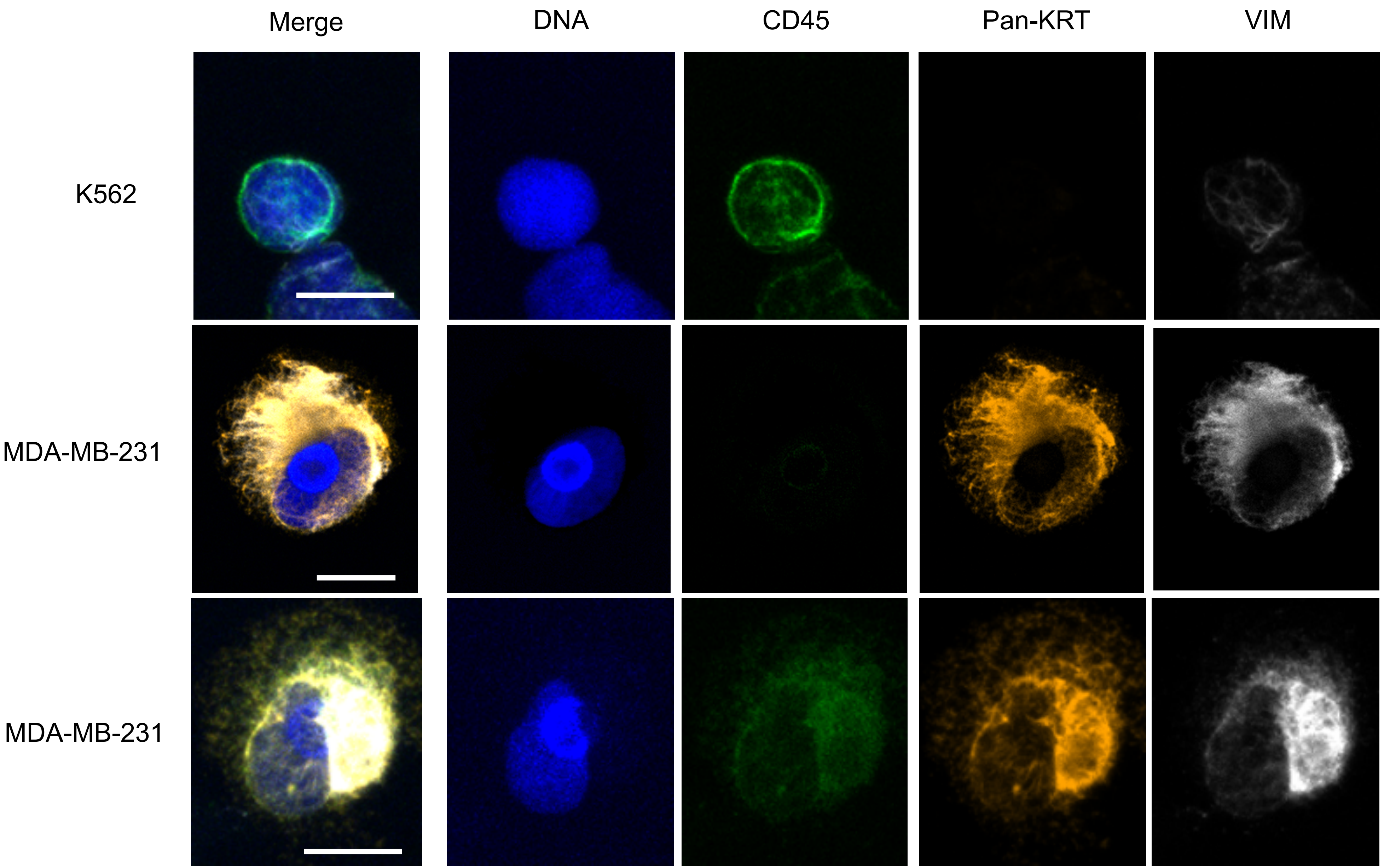

**Supplementary Figure 2c**

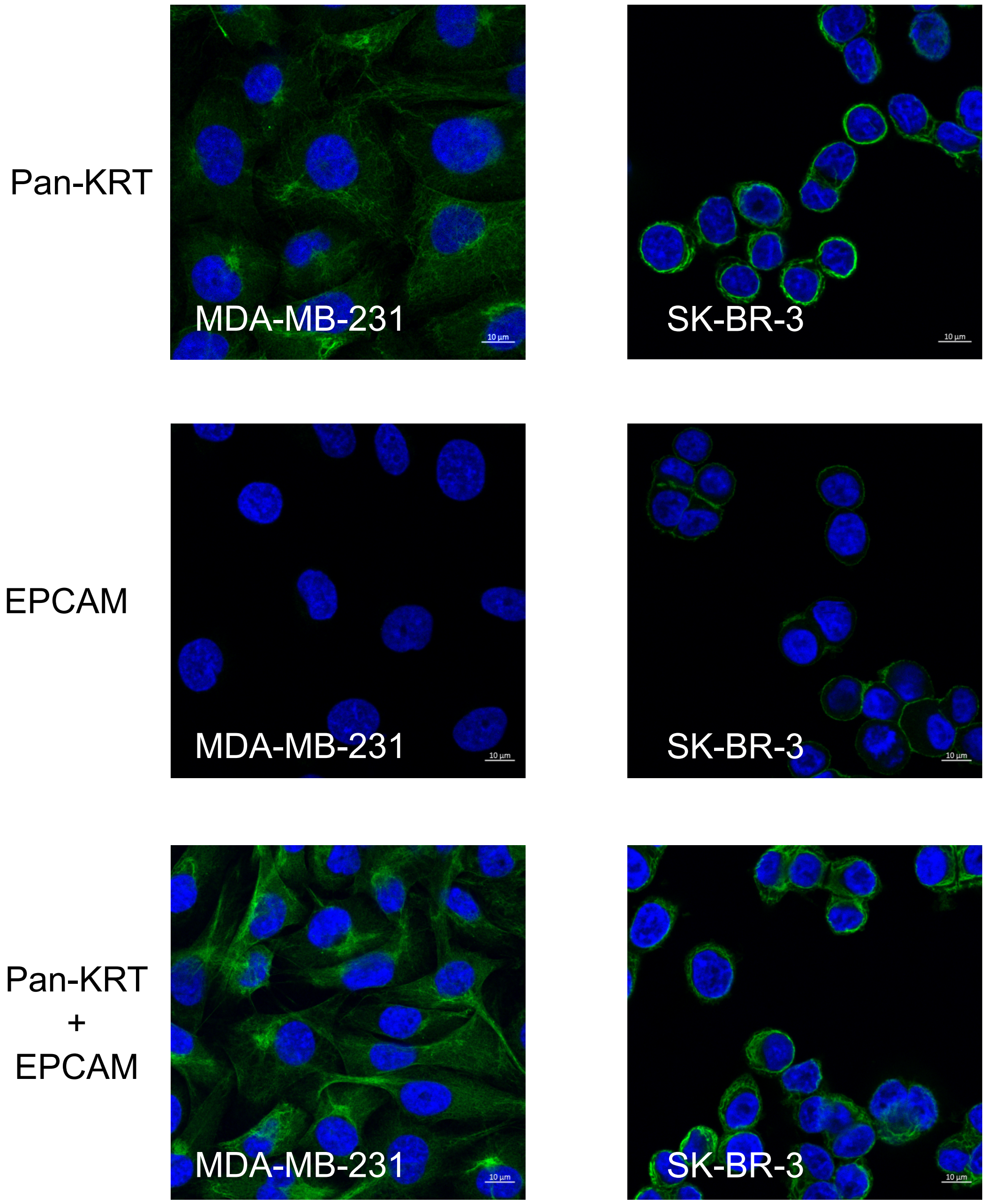

**Figure S2: Antibody validations**

**a. Antibodies specificity**

Each antibody was tested on positive and negative control cells by immunofluorescence to assess their specificity and the signal/noise ratio. Pan-Cytokeratin showed a cytoplasmic staining in MDA-MB-231 (and SK-BR-3 not shown) cells. It was more or less filamentous depending on expression levels and cell spreading. Hs 578t cells were negative for cytokeratin staining. VIM antibody stained the cytoplasm of MDA-MB-231 cells but not SK-BR-3. Anti-EPCAM antibody stained the plasma membrane of SK-BR-3 cells but not MDA-MB-231.CD45 stained the plasma membrane of hematopoietic malignant cells, K-562, but not epithelial cells, MDA-MB-231. ABCB1 antibody was positive on Caco2 cells, with a predominant localization at the membrane (https://www.proteinatlas.org/ENSG00000085563-ABCB1) and negative on MDA-MB-231 cells. LGR5 was detected in a fraction of SUM190 cells, but not on MDA-MB-231 cells. LGR5 shows a dotted intracellular staining (Golgi apparatus and nucleoplasm) in agreement with data validated by the human protein atlas website (https://www.proteinatlas.org/ENSG00000139292-LGR5). LGR5 is not detected in MDA-MB231 cells. Of note, and as expected, markers staining's in cell lines were always stronger than what we could observe in the majority of cells isolated from patients in our study.

**Référence**

Surre J, Saint-Ruf C, Collin V, Orenga S, Ramjeet M, Matic I. Strong increase in the autofluorescence of cells signals struggle for survival. Sci Rep. 2018 Aug 14;8(1):12088.

Supplementary Figure 3

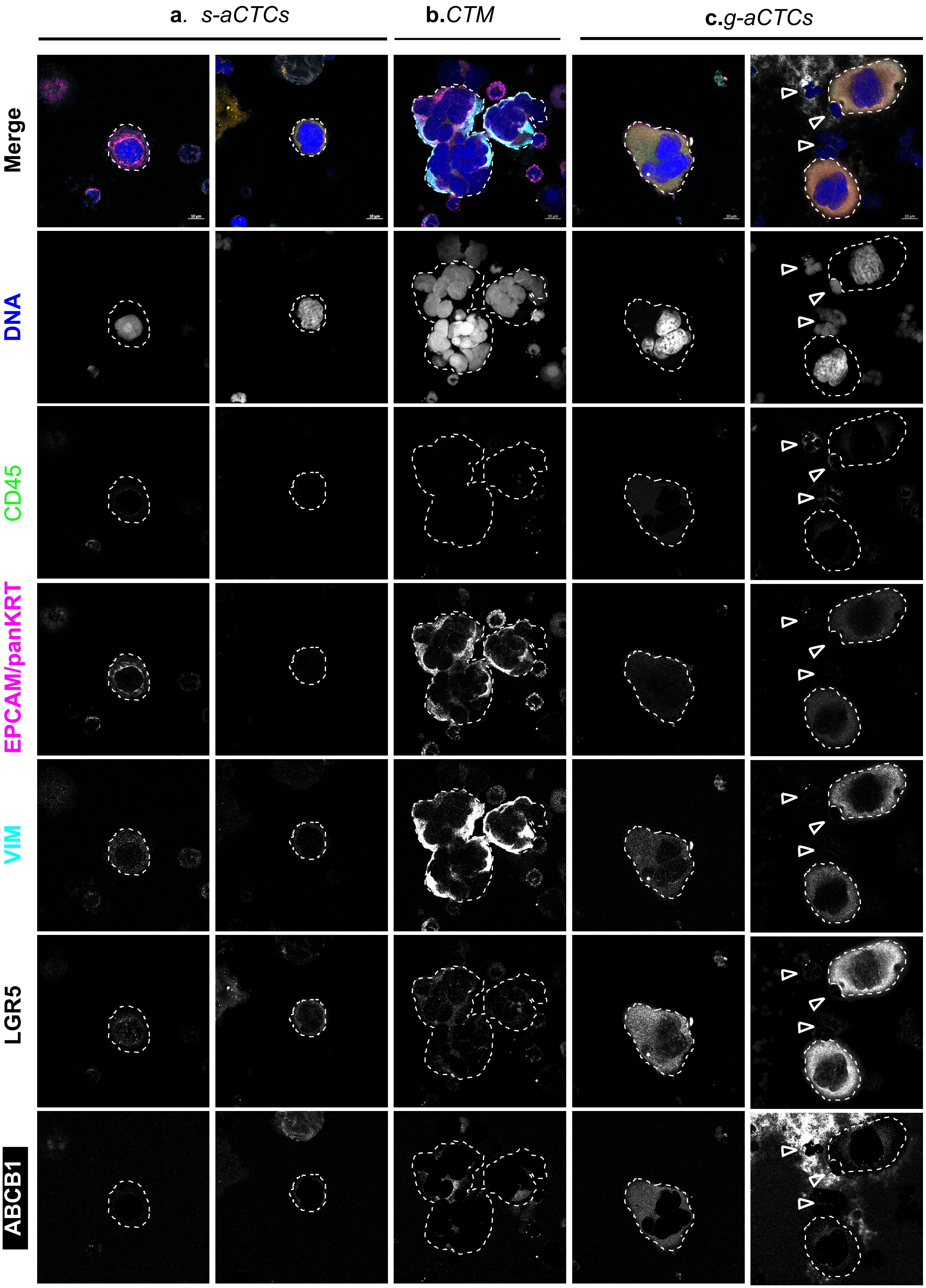

**Figure S3: Examples of the immunofluorescence staining of atypical cells isolated on ScreenCell® filters.**

The blood of mBC patients were filtered and cells on the filter were immunostained. Blood cells are detected with the expression of CD45 antigen at the plasma membrane (empty arrowheads), whereas *aCTCs* (marked with dotted lines) are detected *via* the expression of epithelial markers (EPCAM and pan-KRT) and mesenchymal marker (VIM). Stem cell marker (LGR5) and drug resistance marker (ABCB1) expression are also assessed. Representative images of **a.** *s-aCTCs:* CD45-, EPCAM/Pan-KRT-, VIM+, LGR5+, ABCB1+, and CD45-, EPCAM/Pan-KRT-, VIM+, Lgr5+, ABCB1-, **b.** *CTM:* CD45-, EPCAM/Pan-KRT+, VIM+, LGR5+, ABCB1+, and **c.** *g-aCTCs*: CD45 diffuse, EPCAM/Pan-KRT+, VIM+, LGR5+, ABCB1+, and CD45-, EPCAM/Pan-KRT+, VIM+, LGR5+, ABCB1-, are shown and indicated with dotted lines) Empty arrow heads point blood cells (CD45 positive staining the cell surface).  Scale bar represents 10µm.

### Supplementary Figure 4

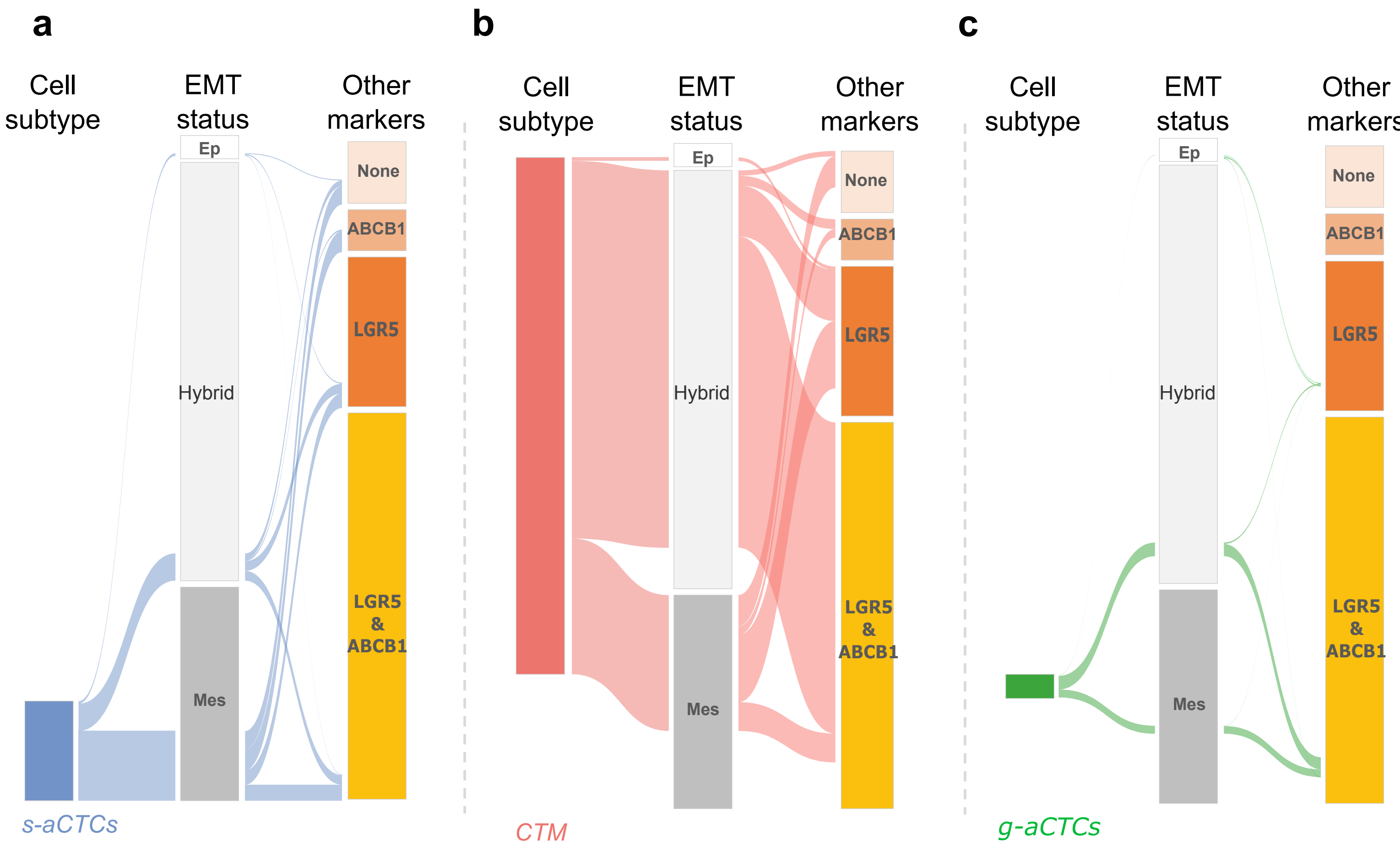

**FigureS4: Alluvial plot representation of the combined expression of EMT, stem and drug resistance markers on aCTCs subsets individually (*CTM, g-aCTCs*, and *s-aCTCs*).** The same data as in figure 3 are represented for each cell subset separately. Alluvial plot representation of the correlations between *aCTC* subsets and molecular markers. The graph shows for the three subsets of atypical cells separately (for *s-aCTCs* in **a***,* for *CTM* in **b** and for *g-aCTCs* in **c**) the combined expression of EMT, LGR and ABCB1 markers. The height of the blocks represents the size of the population. The thickness of a stream represents the number of cells contained in blocks interconnected by the stream. EMT status: Ep=epithelial, Hybrid=epithelial + mesenchymal, Mes=mesenchymal; Other markers: ABCB1, LGR5, Mixed= ABCB1 + LGR5.

### Supplementary Figure 5

a

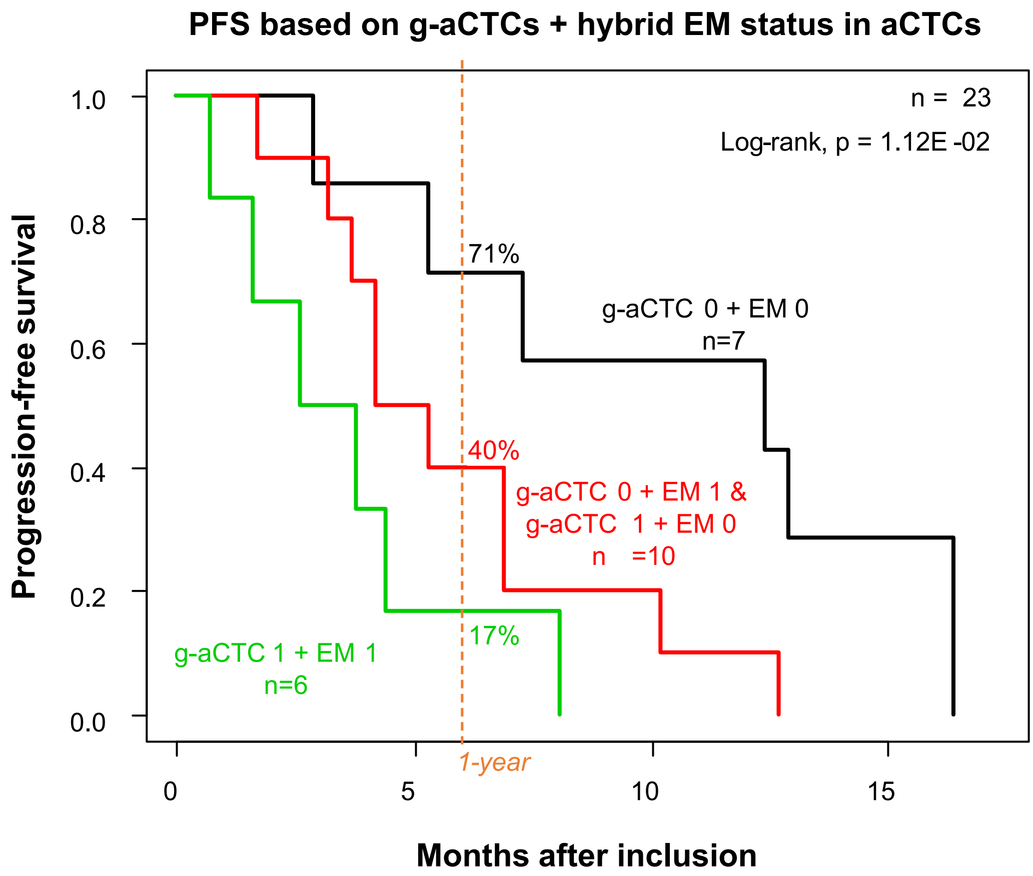


b

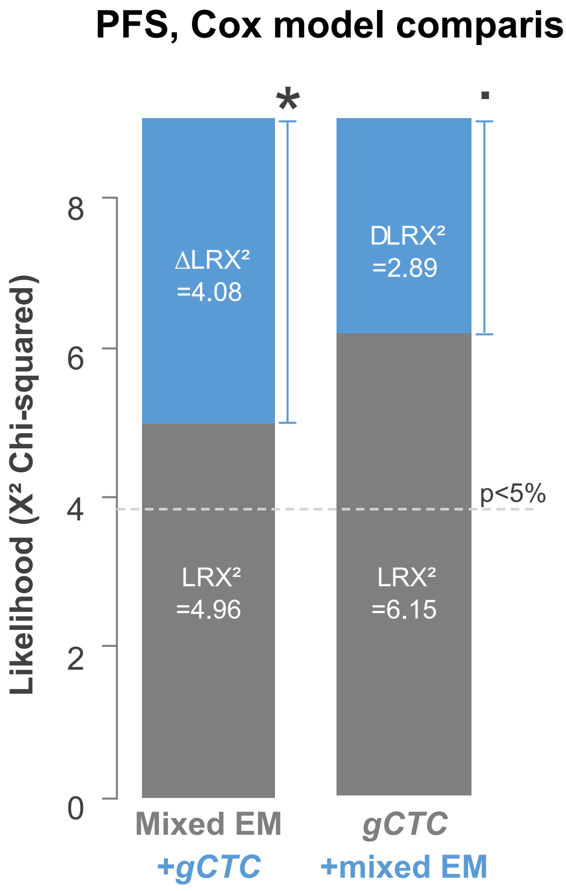

**Figure S5: Kaplan-Meier curves of PFS of patients based on the positivity of *g-aCTCs* and intermediate EM status.**

**a** Survivals were calculated using the Kaplan-Meier method and were compared with the log-rank test to evaluate the prognostic value of combined *g-aCTC* and intermediate EM statuses. Presence of *g-aCTC* is marked as 1 vs absence as 0. Presence of an intermediate EM status in all aCTCs' subsets is represented as 1 value, absence as 0. **b** Prognostic complementarity: the values are given for prognostic information of each variable colored in grey (Mixed EM and *g-aCTC*) on its own (LR-χ2) and when added to the other variable colored in blue (ΔLR-χ2). * indicates $p \leq 0.05$ and indicates trend for significance with $p \leq 0.10$.

### Supplementary Figure 6

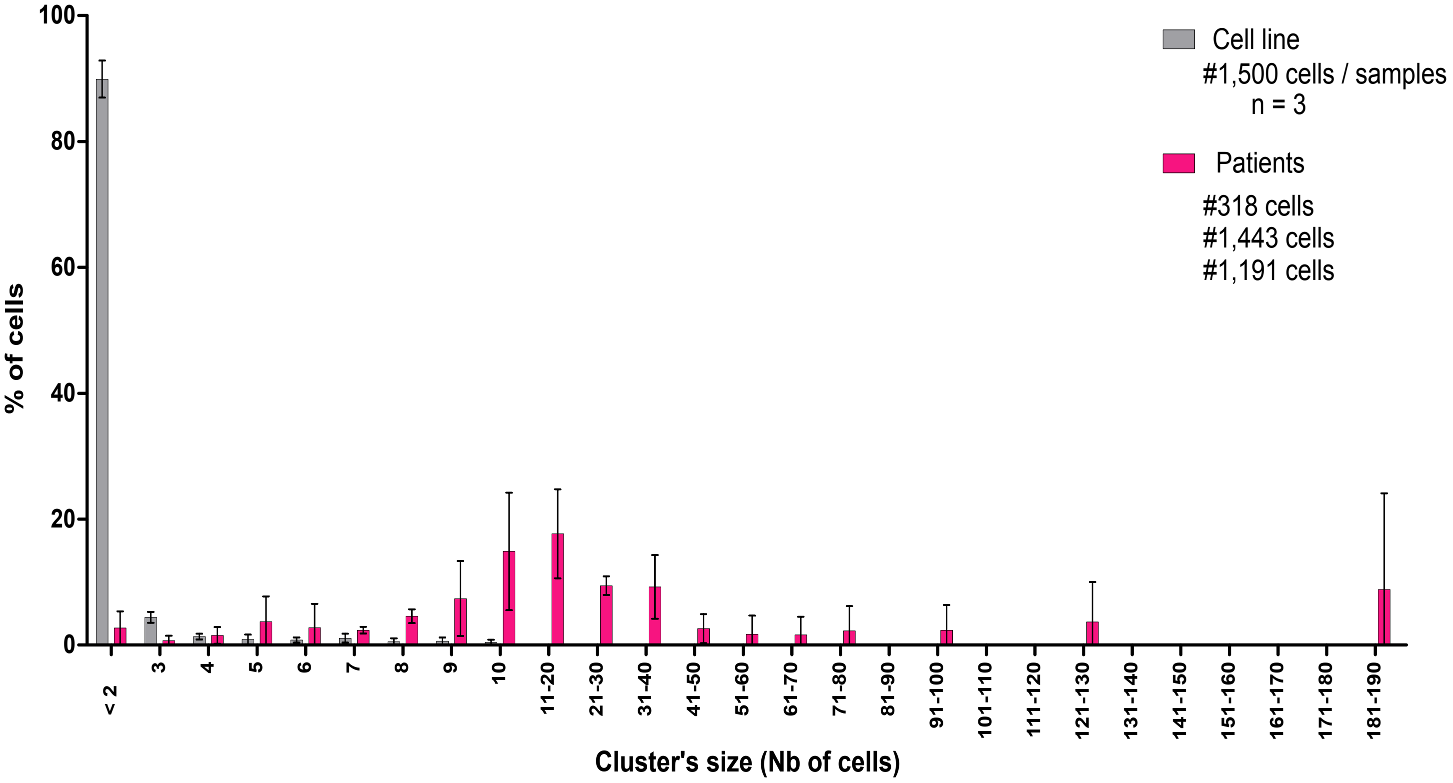

**Figure S6: Clusters verification**

To address potential artificial formation of clusters during the filtration process 1,500 “single” cells (which corresponds to the highest concentration of *a-CTCs* observed in patient) from a cell line SK-BR-3 or MDA-MD-231 (trypsinized and counted) were mixed into 3 ml of blood. The blood was then process as usually within 4 hours with the ScreenCell®Cyto module and stained with May Grundwald Giemsa to count clusters. The results are compared with cluster occurrence in patient samples containing an equivalent number of *a-CTCs* (318, 1,191 and 1,443 *a-CTCs* in 3mls of blood).
